# Climatic suitability for leishmaniasis at global and European scales

**DOI:** 10.64898/2026.05.19.26353551

**Authors:** Gina E C Charnley

## Abstract

Leishmaniasis, a climate-sensitive zoonotic neglected tropical disease, is transmitted by Phlebotomine sand flies and closely linked to socio-economic inequities. Understanding its spatio-temporal dynamics under environmental and social change is critical for effective control. A machine learning framework (XGBoost) was developed to map the global and European distribution of leishmaniasis, incorporating climatic indicators, land cover, elevation, and socio-economic indices (Human Development Index, AROPE). For Europe, five proven vector species (*Phlebotomus perniciosus, P. ariasi, P. perfiliewi, P. neglectus,* and *P. tobbi*) were modelled alongside cutaneous and visceral leishmaniasis. Across both analyses, land use features, particularly shrubland and forest cover, had the greatest explanatory power, reflecting their role in providing microclimates and vertebrate hosts for sand flies. Climatic factors, notably mean temperature of the coldest quarter and humidity of the warmest/driest quarters, were also influential, as these facilitate sand fly survival. Socio-economic predictors consistently improved model performance, confirming the role of poverty and inequity as determinants of disease distribution. Globally, leishmaniasis risk increased by ∼17% since the 1990s, with Africa, Asia, and the Americas experiencing the greatest rise. In Europe, modest continental-scale increases (CL +1.28%; VL +2.47%) masked strong sub-national heterogeneity, including northward expansion of visceral leishmaniasis and increases in cutaneous leishmaniasis in southern and eastern regions. Sand fly projections indicated expansion of warm-adapted species (*P. ariasi, P. perniciosus, P. neglectus*) and contraction of species preferring cooler, more humid niches (*P. perfiliewi, P. tobbi*). These findings highlight climate change, land use, and inequity as interacting drivers of leishmaniasis, emphasising the need for enhanced surveillance, integrated vector management, and targeted support for vulnerable populations, including refugees and migrants.

## Introduction

The leishmaniases (visceral leishmaniasis (VL) and cutaneous leishmaniasis (CL), including mucocutaneous) are parasitic diseases, with a broad spectrum of clinical signs ranging from simple to severe cutaneous ulcers, disfigurement and stigma including destruction of the oral-pharyngeal mucosa, to life-threatening visceral involvement with a case fatality rate of >95% if not treated. The disease is caused by protozoa of the *Leishmania* genus and are transmitted by bites of infected female sand flies of the subfamily Phlebotominae.^1^

Human leishmaniasis is endemic in 99 countries and territories, with an estimated 700,000 to 1 million new human cases reported every year, although this is likely an underestimate. The disease affects some of the world’s poorest and marginalised/displaced people and is associated with malnutrition, migration, poor housing, immunocompromised conditions e.g. HIV co-infection, and lack of financial resources.^2,3^ The highest burden of leishmaniasis is concentrated in only twelve countries, reporting over 90% of cases, these include India, Bangladesh, Sudan, South Sudan, Ethiopia, and Brazil for VL and Afghanistan, Algeria, Brazil, Colombia, Ethiopia, Iran, Peru, and Syria for CL.^4^

There are around 30 species of sand fly known to transmit the disease to humans and over 20 species of *Leishmania* parasites which cause disease in humans, including *L. donovani, L. infantum*, and *L. major,* which cause VL, and species like *L. tropica*, *L. amazonensis*, and *L. braziliensis*, responsible for CL. Species of the disease and main vectors are generally associated with transmission in specific geographic areas such as sand fly species *Phlebotomus argentipes* in the Indian subcontinent and *Lutzomyia longipalpis* in Latin America (see **Fig 1**).^5^

**Fig 1.**
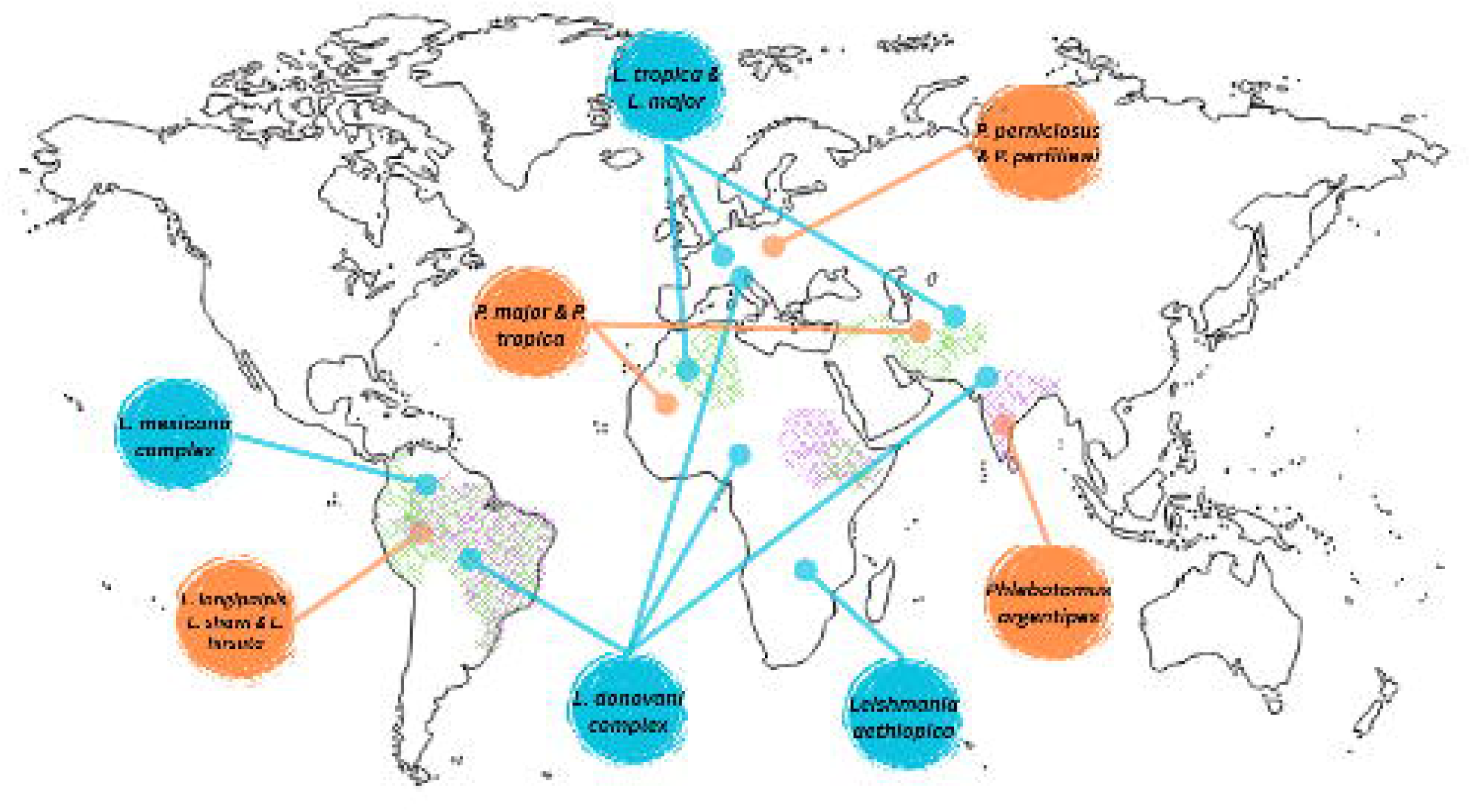
Distribution of leishmaniasis burden globally, the hatching shows where >90% of leishmaniasis cases are reported, with visceral in purple and cutaneous in green. Additionally, the blue circles highlight some of the main parasites responsible for disease in specific areas and the orange circles, the main vector species for transmission.^4,5^

The leishmaniases are climate-sensitive zoonotic diseases which are distributed between latitude 50° N and latitude 40° S,^5^ with a wide variety of vertebrate reservoir hosts and blood sources, including humans, domestic dogs, lagomorphs, and synanthropic and wild rodents. Sand fly species distributions tend to be restricted to warmer areas, although optimum temperature and humidity conditions for sand fly activity vary substantially between species and regions, influencing sand fly metabolism, and the development and survival of the sand fly life stages.^6–9^ Humidity and soil moisture are important variables for egg survival, whereas heavy rainfall can destroy suitable breeding sites and/or kill immature stages.^10^

Temperature appears to be one of the main drivers enabling spatial range expansions of many vectors into higher latitudes and altitudes under climate change^11–15^. Warmer temperatures are expected to reduce the time required for *Leishmania* parasites to undergo biological development into the infectious form inside the vector, such that an infected vector is infectious for longer periods in relation to its life expectancy. Conversely, high temperatures may variably reduce the lifespan of some vectors which can reduce their infectious period for onward transmission. Projecting sand fly populations under climate change scenarios, accounting for landscape features, confirms a vector geographical expansion into new ecological niches from where they are currently present^12,16–18^. However, there are several anthropogenic factors which may alter this, including poverty alleviation and land use changes.

Leishmaniasis is considered an emerging threat in Europe, especially due to climate change. VL is already endemic in a number of countries in the Balkans and Mediterranean region (including Spain, Italy, Portugal, France, Greece, Turkey and Albania), and CL is considered an emerging threat. Cold tolerant species of sand fly (e.g. *P. mascittii*, and *P. perniciosus*) are present in northern France, parts of Switzerland, Austria, Germany and Belgium, all regions where *Leishmania* is currently non-endemic.^19,20^ However, the risk of establishing local transmission of *L. infantum* in non-endemic regions where competent vectors are present following the introduction of infected dogs is considered high (47%-72%): This is proposed to have led to the emergence of new disease foci in northern Italy and northern Spain.^21–24^ Furthermore, the Madrid leishmaniasis outbreak starting in 2009 found hares and rabbits (abundant in many parts of Europe) in peri-urban green spaces to be the main reservoirs for infection.^25^

There is no human vaccine for leishmaniasis, and standard chemotherapeutic treatments are often highly toxic, costly and access and compliance to treatment can be low, therefore, prevention is currently the best option for disease control. In the absence of reliable surveillance data of disease presence, epidemiological modelling is helping to better delineate areas where insecticide treatments for humans, dogs and the environment may be needed and identify areas where there is a high prevalence of asymptomatic human and zoonotic carriers.^26–28^ In continuing these efforts, here a machine learning approach was used with the aim to map the distribution of leishmaniasis based on the climatic suitability of the vector and socio-economic risk factors which lead to disease. The analysis provides a novel contribution, performing the analysis at both a global and regional scale (Europe), mapping the historical risk in both endemic and emerging regions against a range of key risk factors.

## Methods

### Data sources and harmonisation

Leishmaniasis data were extracted from Pigott *et al.* (2014)^29,30^ which aimed to estimate the global distribution of leishmaniasis, and included a case point (x,y) dataset at an annual temporal scale. Due to differences in reporting, data processing differed for the global and European analysis. Cases were transformed into a binary presence/absence outcome variable (leishmaniasis occurrence) and for the global analysis attributed to the national level as a combined CL & VL outcome, whereas, for the European analysis, cases were to the NUTS2 level and CL and VL kept as separate outcomes (due to a greater availability of fine-scale data). Models were fit to the full duration of the dataset 1970-2012, which was used as the baseline period, and the World Health Organization Global Health Observatory data used as validation for near-term predictions in the national global analysis (2005-2024).^31^ The spatial distribution of the leishmaniasis data used as the outcome in model training are shown in **S1 Fig**.

There are no global sand fly datasets currently available, therefore the inclusion of such data was limited to the European analysis, with the global analysis, the assumption was made that the presence of the disease meant the presence of the vector, using leishmaniasis data as a proxy for sand fly presence. The data were extracted from the European Centre for Disease Prevention and Control (ECDC) annual sand fly presence data via VectorNet, these data were to the point level (x,y) and extrapolated to the NUTS2 level as a binary outcome (presence/absence) for the full dataset length 2015-2024 (see **S2 Fig** for presence and absence for 2024).^32^ In Europe, there are many endemic sand fly species, however, not all of them are proven or suspected vectors of *L. infantum* (the main causative agent of leishmaniasis in Europe). Therefore, the European sand fly data were subset to species which are proven leishmaniasis vectors, namely: *Phlebotomus. perniciosus*, *P. ariasi*, *P. perfiliewi*, *P. neglectus*, and *P. tobbi*.^33^

Two-meter air temperature, dew point temperature, volumetric soil water and precipitation data were extracted from ERA5 monthly re-analysis via the Copernicus Climate Data Store for the full time-series (1951-2025) at a 0.25°x0.25° grid cell resolution.^34^ Two-meter temperature (in Kelvin) and 2-meter dew point temperature (in Kelvin) were transformed into degrees Celsius (value in K - 273.15). Using temperature and dew point temperature, relative humidity was calculated using the August-Roche-Magnus equation,^35^

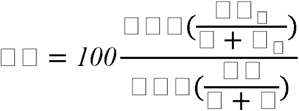

where *a* and *b* are constants of 17.625 and 243.04, respectively and *T_d_* refers to dew point temperature and *T* to air temperature. Using the monthly climate data, the variables were then converted to the WorldClim annual bioclimatic indicators, these include nineteen variables to represent Essential Climate Variables relevant to the biodiversity community (see **Table 1** summary and the full explanations and equations in **S1 Table**).^36^ The nineteen variables originally only use temperature and precipitation data, however, here these principles were expanded to humidity and soil water data, which are important for sandfly development and breeding. The climate data were then extrapolated to national/NUTS2 averages to match the spatial scale of the outcome for the two spatial scales.

**Table 1:**
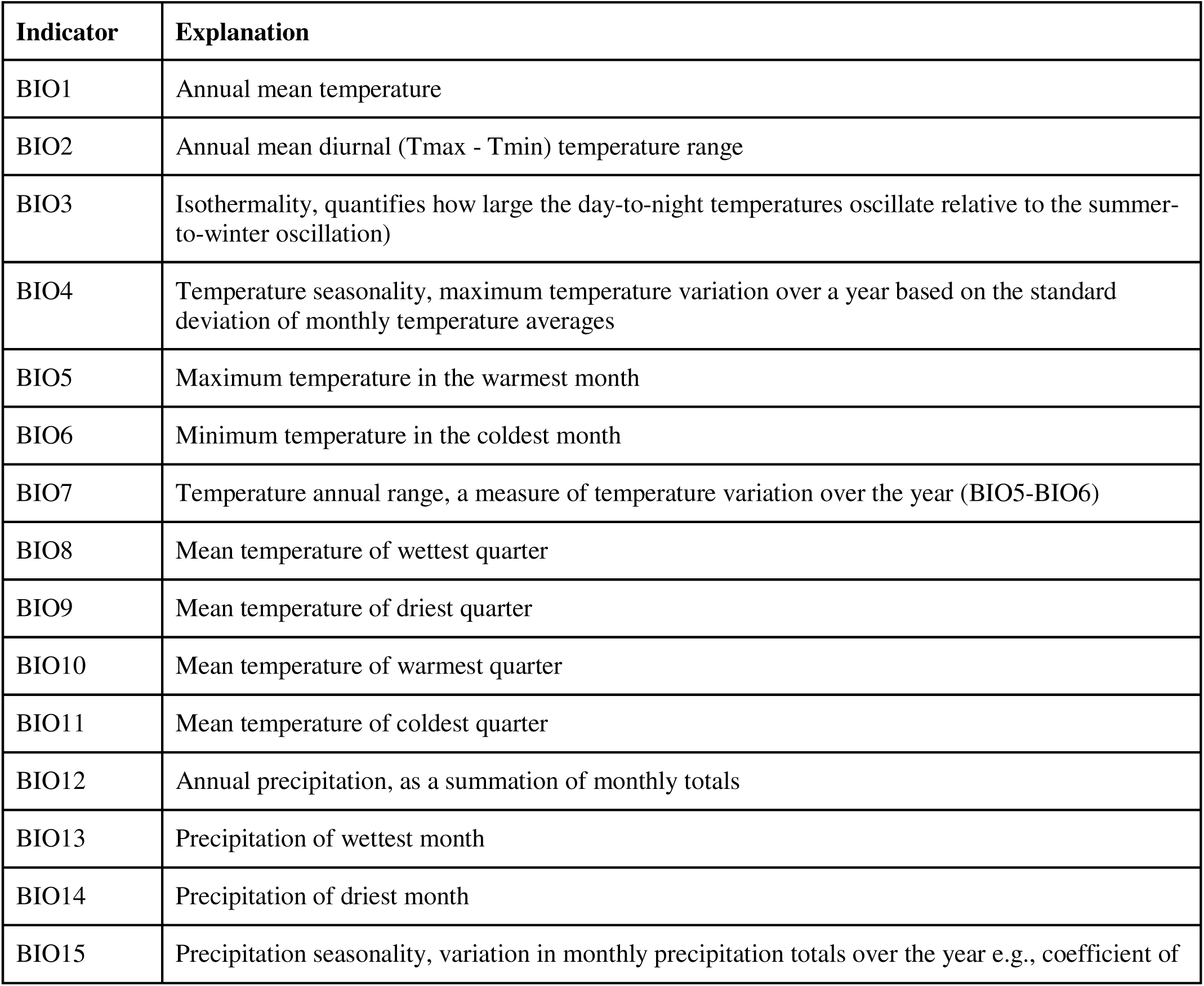
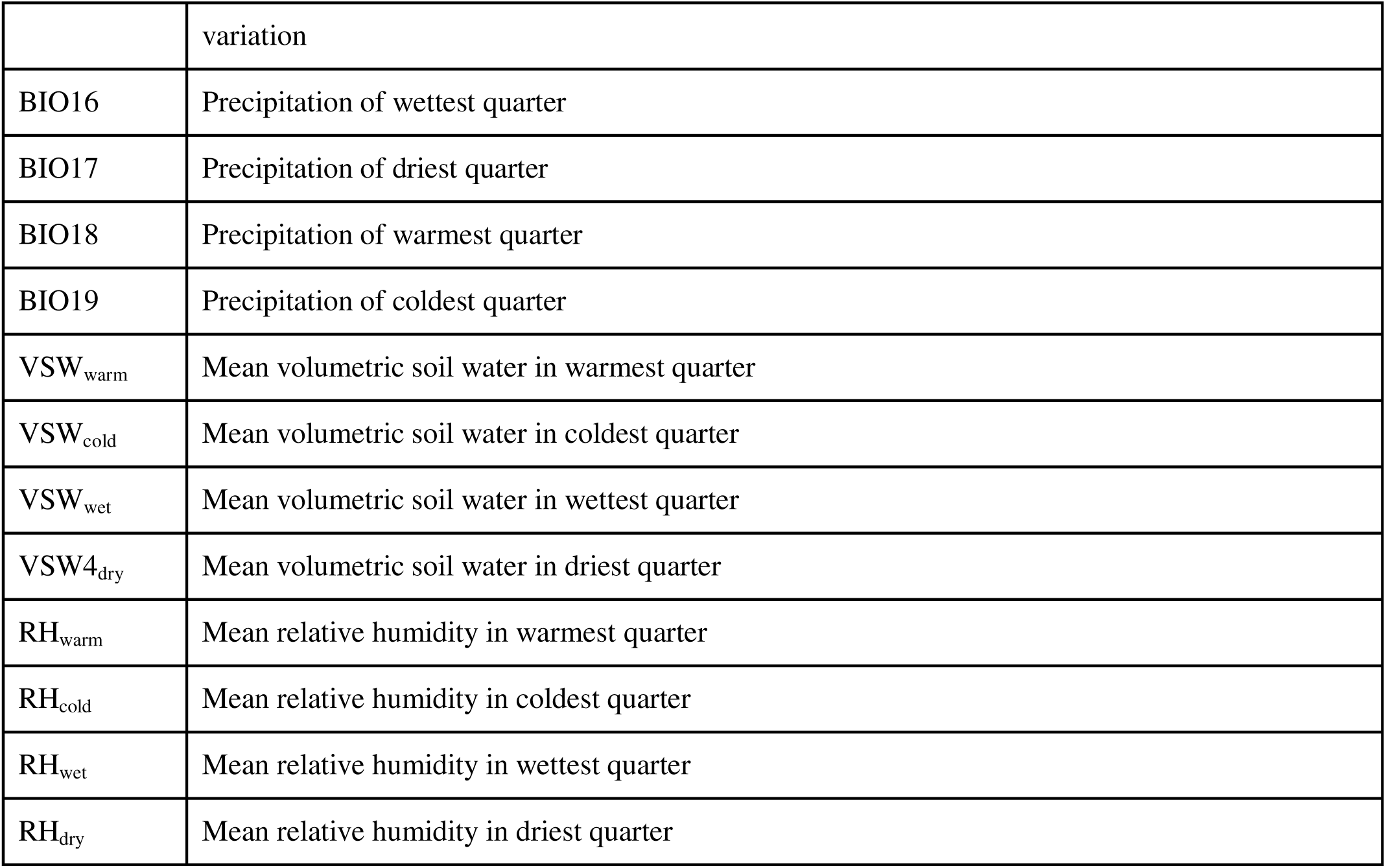
Summary of the twenty-seven climate covariates considered in the modelling approach. Quarters are defined as Jan-Mar, Apr-Jun, Jul-Sept & Oct-Dec. ^36^

| Indicator | Explanation |
| --- | --- |
| BIO1 | Annual mean temperature |
| BIO2 | Annual mean diurnal (Tmax - Tmin) temperature range |
| BIO3 | Isothermality, quantifies how large the day-to-night temperatures oscillate relative to the summer-to-winter oscillation) |
| BIO4 | Temperature seasonality, maximum temperature variation over a year based on the standard deviation of monthly temperature averages |
| BIO5 | Maximum temperature in the warmest month |
| BIO6 | Minimum temperature in the coldest month |
| BIO7 | Temperature annual range, a measure of temperature variation over the year (BIO5-BIO6) |
| BIO8 | Mean temperature of wettest quarter |
| BIO9 | Mean temperature of driest quarter |
| BIO10 | Mean temperature of warmest quarter |
| BIO11 | Mean temperature of coldest quarter |
| BIO12 | Annual precipitation, as a summation of monthly totals |
| BIO13 | Precipitation of wettest month |
| BIO14 | Precipitation of driest month |
| BIO15 | Precipitation seasonality, variation in monthly precipitation totals over the year e.g., coefficient of |
|  | variation |
| BIO16 | Precipitation of wettest quarter |
| BIO17 | Precipitation of driest quarter |
| BIO18 | Precipitation of warmest quarter |
| BIO19 | Precipitation of coldest quarter |
| VSW<sub>warm</sub> | Mean volumetric soil water in warmest quarter |
| VSW<sub>cold</sub> | Mean volumetric soil water in coldest quarter |
| VSW<sub>wet</sub> | Mean volumetric soil water in wettest quarter |
| VSW<sub>dry</sub> | Mean volumetric soil water in driest quarter |
| RH<sub>warm</sub> | Mean relative humidity in warmest quarter |
| RH<sub>cold</sub> | Mean relative humidity in coldest quarter |
| RH<sub>wet</sub> | Mean relative humidity in wettest quarter |
| RH<sub>dry</sub> | Mean relative humidity in driest quarter |

Land cover data were from the most recent (2019) Copernicus Land Monitoring Service’s Global Land Cover raster files at 100m resolution.^37^ To harmonise spatial scales to the required outcome (∼29km grid), the land cover data were re-sampled using bilinear interpolation (as the variable is continuous and requiring a smooth interpolation). The resulting data were the percentage of land cover by each Copernicus land class: bare, crops, grass, moss/lichen, shrub, snow, forest/tree, urban/built-up, permanent and seasonal water, at a 0.25°x0.25°.

To improve the model’s predictive performance, additional environmental and socioeconomic conditions were included, such as NASA’s Shuttle Radar Topography Mission Global elevation data,^38^ NASA’s Gridded Population of the World,^39^ and the United Nations Human Development Index^40^ for the global analysis and EUROSTAT’s at-risk of poverty and social exclusion (AROPE)^41^ for the Europe-level analysis. Sand fly survival is known to decrease with increasing elevation due to less suitable climates,^12^ while leishmaniasis is considered a disease of inequity, and is linked to poorer communities with less access to healthcare.^42^ The harmonised datasets consist of 7,103 (5,328 training and 1,775 testing) observations at the global scale and 15,500 (11,625 training and 3,875 testing) at the European level for the 1970-2012 training period.

### Preliminary models

A machine learning approach was used, looking to advance previous modelling exercises that were limited in spatial scale, temporal resolution and number of predictors,^43^ to predict the risk for leishmaniasis according to climatic suitability for the vectors and the socio-economic risks associated with the disease, in the training period 1970-2012 and subsequently generated annual predictions for 1951-2024 (creating both in sample and out-of-sample predictions). The model was trained on a variety of climatic and socioeconomic features against a binary leishmaniasis and sand fly absence/presence outcome variables, which varied depending on data availability at the two spatial scales.

The extreme gradient boosted regression (XGBoost) algorithm was used here and is a machine learning algorithm that creates an ensemble of weak decision trees to form a stronger prediction model by iteratively learning from weaker classifiers and adding them to a strong classifier (boosting). Gradient boosted regression is a flexible methodology that allows for non-linearity, both among features (interactions) and between features and the outcome, collinearity between features and non-random patterns of missing data. Additionally, XGBoost allows the use of regularisation parameters to prevent overfitting models to small, unbalanced datasets.^44^

First, a preliminary set of models were ran with the full set of features against each of the 7 outcomes, for the global analysis, leishmaniasis occurrence and for the European analysis, the presence of *P. perniciosus*, *P. ariasi*, *P. perfiliewi*, *P. neglectus*, *P. tobbi*, VL and CL. The preliminary analysis aimed to identify key contribution variables according to the explainable AI framework Shapley Additive Explanations (SHAP),^45^ which uses game theory to fairly distribute the model’s output among the predictions by calculating the average marginal contribution of each feature across all possible subsets, considering feature interactions to improve reliability over impurity-based methods (e.g., Gini Impurity). Additionally, Pearson correlation coefficient values were calculated to evaluate relationships among the 27 climate-related variables (as these were expected to be highly correlated due to them relying on the same underlying weather data), land classes and elevation, clustered at r > 0.7 to avoid overfitting.

### Model training, testing & tuning

Once the best subset (top 10 in terms of Importance, defined as a normalized average improvement in model accuracy when a feature is used in a split) of clustered bioclimatic indicators were selected, these were merged to the socioeconomic data and the dataset was split at 75%:25% training:testing. Model predictive performance was assessed in two stages based on threshold-dependent (*e.g*., sensitivity/specificity) and threshold-independent (*e.g*., AUC) metrics, the first using k-fold cross-validation on the training dataset and the second on the unseen testing dataset, to provide reliable validation and an independent evaluation.

Four models were selected for each outcome and spatial scale for further improvements via hyperparameter tuning, these included the top two models which contained solely environmental features (climate, land use, elevation) and the top two which contained environmental and socio-economic features (population and inequity, if these improved model performance). Model performance was separated to models with and without socioeconomic data to evaluate the potential importance of the environment on leishmaniasis transmission. A variety of different parameter settings were explored (see **Table 2**), with an early stop feature to monitor overfitting. Out-of-sample leishmaniasis data from the WHO Global Health Observatory^1^ were used to assess the performance of the predicted risk for the remaining time-period (2012-2024) at the global scale.

**Table 2:**
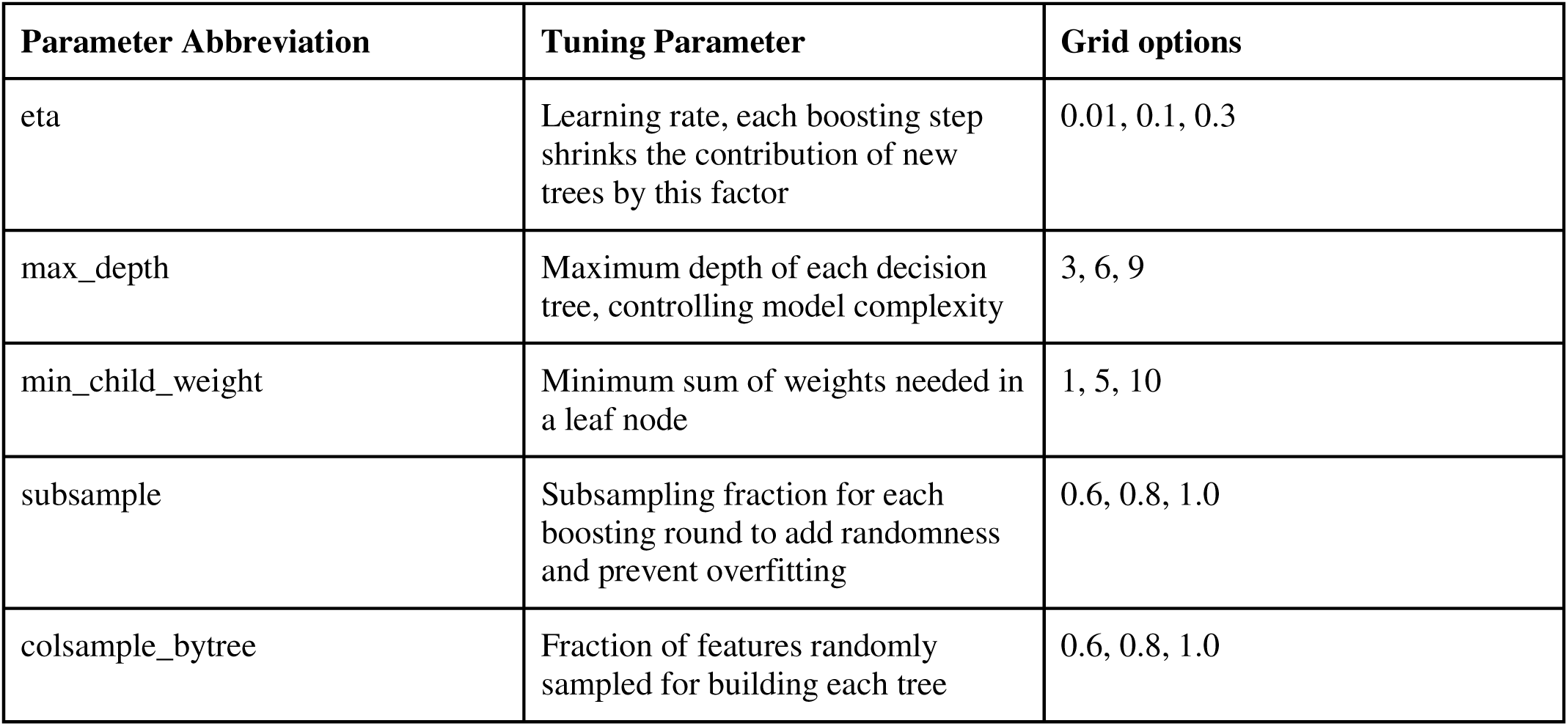
Tuning parameters which were used for the grid search on the subset of best models.

| Parameter Abbreviation | Tuning Parameter | Grid options |
| --- | --- | --- |
| eta | Learning rate, each boosting step shrinks the contribution of new trees by this factor | 0.01, 0.1, 0.3 |
| max_depth | Maximum depth of each decision tree, controlling model complexity | 3, 6, 9 |
| min_child_weight | Minimum sum of weights needed in a leaf node | 1, 5, 10 |
| subsample | Subsampling fraction for each boosting round to add randomness and prevent overfitting | 0.6, 0.8, 1.0 |
| colsample_bytree | Fraction of features randomly sampled for building each tree | 0.6, 0.8, 1.0 |

### Model predictions

The model with the best performance was used to predict risk of leishmaniasis for 1951-2024 at an annual national scale at the global level and at the annual and NUTS2 levels for Europe. Furthermore, the models with the best predictive performance for each fly species were used to make annual predictions of sand fly presence risk from the start of the full training period (1970-2024) for each NUTS2 region in Europe. After extracting the predictions for the sand flies, these were joined to the prediction dataset, and four models were used to predict leishmaniasis risk, two using solely the best fit leishmaniasis models, and two using the best fit models and the fly predictions as features. Four leishmaniasis models were trained to understand how adding the sand fly predictions as features altered the leishmaniasis predictions. A schematic of the full modelling approach for the two spatial scales is shown below in **Fig 2**.

**Fig 2.**
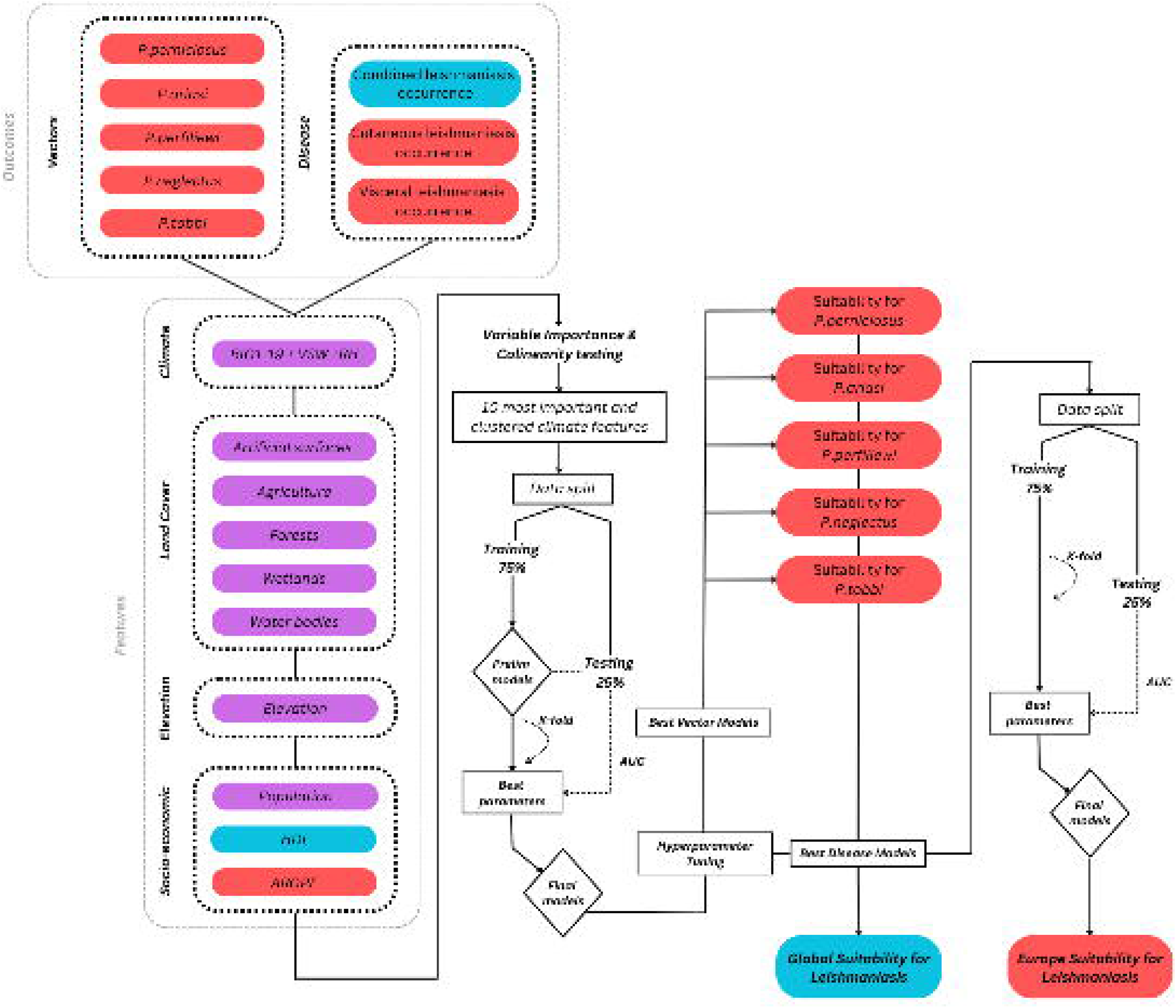
Schematic of the modelling approach used here, including covariate exploration and threshold-dependent and threshold-independent model performance on the training and testing data. Data input and stages which were only part of the global analysis are in blue, only part of the Europe analysis in red and used in both are in purple and white.

## Results

For the leishmaniasis data used in this study, there were several spatial and temporal patterns and heterogeneities. As with most disease datasets, more cases were reported in the near-term, with a general increase in the number of reported cases after the 1980s and particularly after 2000. These reflect global efforts in disease reporting since the millennium and an increase in reporting infrastructure and innovation. Spatially, the most cases were reported in South America, North Africa, South and Southwest Asia and southern Europe. The distribution generally reflects the global burden of leishmaniasis, with a particularly high number of cases reported in Brazil and India (see **S1 Fig**).

After extracting the SHAP values, the top ten climate-related features were selected for consideration in the best-fit model via each unique combination of formulas, selecting only one feature per cluster to prevent overfitting. Across both scales (global and European) and outcome variables (leishmaniasis and sand flies) several of the same features were repeatedly selected and had high SHAP values. Land use classes provide the most powerful predictors, namely shrubs, the presence of waterbodies, urban areas, grass and forests, and for the European analysis, the AROPE inequity index. In terms of the climatic variables, BIO11 (mean temperature in the coldest quarter) was selected across five of the best fit models, along with humidity in the warmest and driest quarters having relatively high SHAP importance values (see **Fig 3** and for the full SHAP, correlation and feature subset results **S3-4 Figs** & **S2 Table**). As expected, several of the land use and bioclimatic indicators were clustered at the correlation coefficient threshold.

**Fig 3.**
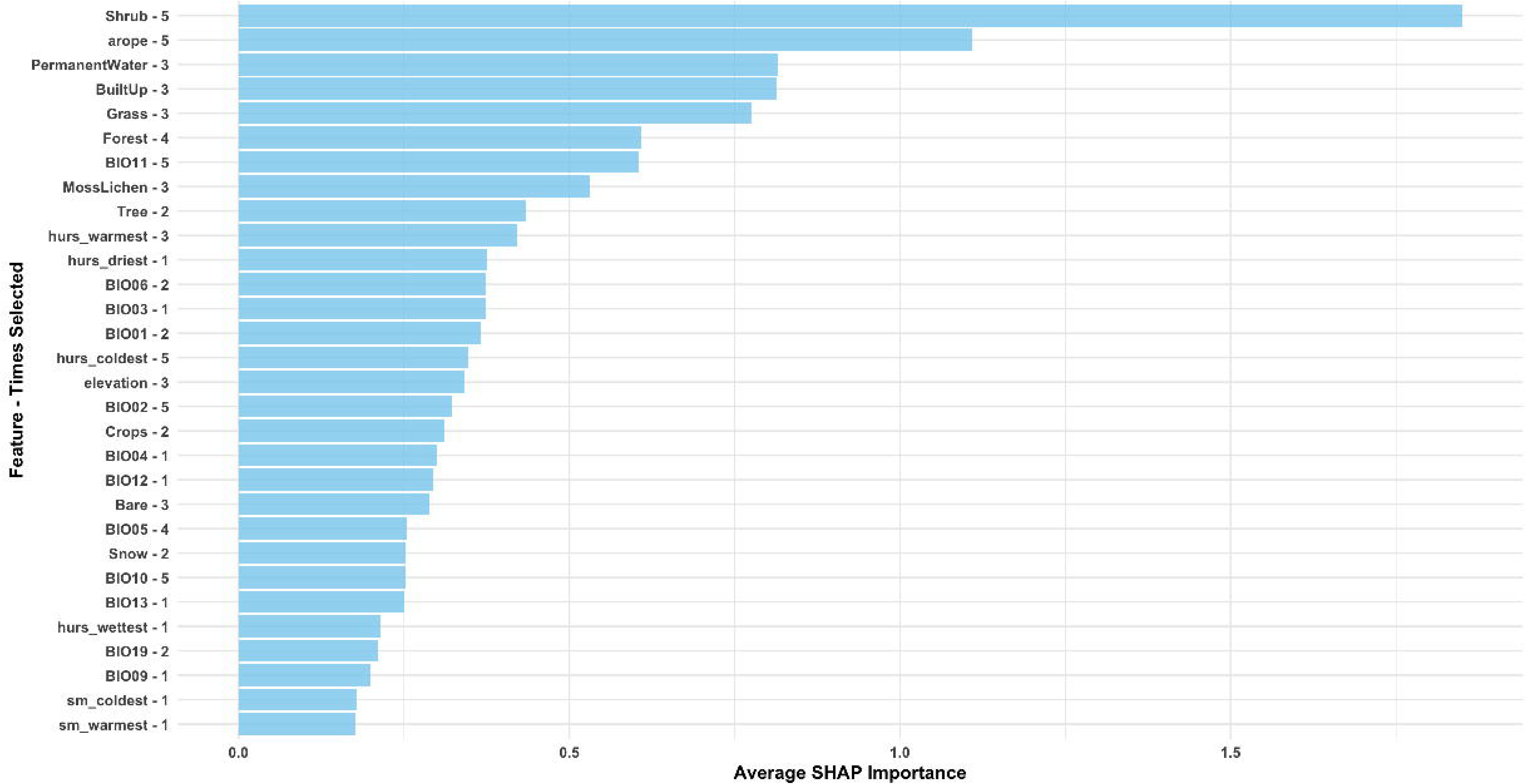
The average SHAP importance values of the top 10 features selected across the eight outcome variables tested here. The y-axis shows the name of the feature, followed by the number of times it was selected in the top 10 subset for the different outcomes (maximum 8).

All models performed well with a high AUC (>0.95), suggesting that a combination of environmental and socio-economic covariates are powerful predictors of both sand fly presence and the disease at global and European scales (**Table 3**). The inclusion of socio-economic predictors at both scales increased model performance and proved valuable predictors, being selected in the best fit models in all but one model (Europe *P. tobbi* model). The European models performed slightly better than the global analysis (high AUC and lower RMSE), likely due to better data availability and being able to complete the analysis on a finer spatial scale. Out-of-sample comparisons between the global model predictions and the WHO Global Health Observatory for 2012 onwards found an r = 0.81 and an AUC of 0.72. The inclusion of the sand flies as predictors in the European models created similar performing models in terms of AUC and RMSE, and the predictions followed the same trends, but the inclusion of the flies predicted higher magnitudes of leishmaniasis risk.

**Table 3:** Best fit model performance metrics. RMSE = Root Mean Square Error. AUC = Area Under the receiver operator Curve. Performance metrics are rounded to three decimal places.

| Region | Outcome | Features | Tuning parameters | RMSE | AUC |
| --- | --- | --- | --- | --- | --- |
| Europe | Phlebotomus neglectus | AROPE, shrubs, humidity in the coldest quarter, BIO04, forest, population | eta = 0.3<br>max_depth = 6,<br>min_child_weight = 1<br>subsample = 0.6<br>colsample_bytree = 0.6 | 0.054 | 1.000 |
| Europe | Phlebotomus perfiliewi | AROPE, shrubs, BIO02, BIO11, elevation, forest | eta = 0.1<br>max_depth = 3<br>min_child_weight = 1<br>subsample = 0.6<br>colsample_bytree = 0.6 | 0.108 | 0.999 |
| Europe | Phlebotomus ariasi | AROPE, BIO06, elevation, permanent water, BIO02, forest | ta = 0.3<br>max_depth = 3<br>min_child_weight = 1<br>subsample = 0.6<br>colsample_bytree = 0.6 | 0.024 | 1.000 |
| Europe | Phlebotomus perniciosus | BIO06, AROPE, forest, elevation, grass, population | eta = 0.3<br>max_depth = 9<br>min_child_weight = 5<br>subsample = 1<br>colsample_bytree = 0.8 | 0.031 | 1.000 |
| Europe | Phlebotomus tobbi | BIO02, shrubs, grass, built up, BIO04, crops, forest | eta = 0.3<br>max_depth = 3<br>min_child_weight = 1<br>subsample = 0.6<br>colsample_bytree = 0.6 | 0.031 | 1.000 |
| Europe | Cutaneous leishmaniasis | AROPE, BIO01, elevation, shrubs, humidity in the driest quarter | eta = 0.1<br>max_depth = 9<br>min_child_weight = 1<br>subsample = 0.6<br>colsample_bytree = 0.6 | 0.132 | 0.976 |
| Europe | Visceral leishmaniasis | AROPE, shrubs, elevation, BIO10, humidity in the driest quarter, BIO04 | eta = 0.1<br>max_depth = 9<br>min_child_weight = 5<br>subsample = 1<br>colsample_bytree = 1 | 0.161 | 0.972 |
| Europe | Cutaneous leishmaniasis w/ fly predictions | AROPE, BIO01, elevation, shrubs, humidity in the driest quarter, neglectus, perfiliewi, ariasi, perniciosus, tobbi | eta = 0.1<br>max_depth = 9<br>min_child_weight = 1<br>subsample = 0.6<br>colsample_bytree = 0.6 | 0.131 | 0.975 |
| Europe | Visceral leishmaniasis w/ fly predictions | AROPE, shrubs, elevation, BIO10, humidity in the driest quarter, BIO04, neglectus, perfiliewi, ariasi, perniciosus, tobbi | eta = 0.1<br>max_depth = 9<br>min_child_weight = 5<br>subsample = 1<br>colsample_bytree = 1 | 0.164 | 0.968 |
| Global | Leishmaniasis | Bare, elevation, BIO10, HDI | eta = 0.1<br>max_depth = 9<br>min_child_weight = 10<br>subsample = 0.6 | 0.353 | 0.903 |
|  |  |  | colsample_bytree = 1 |  |  |

The results were compared across several baseline and comparator periods; to highlight changes in risk and the annual results were spatially aggregated by regions to present the full time-series (1951-2024) (see **Fig 4**). Globally, leishmaniasis risk has increased by 17% in 2014-2023, compared to 1990-1999, based on the predictions for the best fit model here. Regions at the greatest risk include Africa, Asia and South and Central America, with a 39%, 34% and 27% predicted risk for 2024, respectively. When sub-setting Asia to solely the Eastern Mediterranean region, this risk for 2024 increased further to 46%. However, changes in predicted risk have only been marginal since the 1990s in the Eastern Mediterranean (8% change), along with Europe (2-6% change) (**Fig 5**).

**Fig 4.**
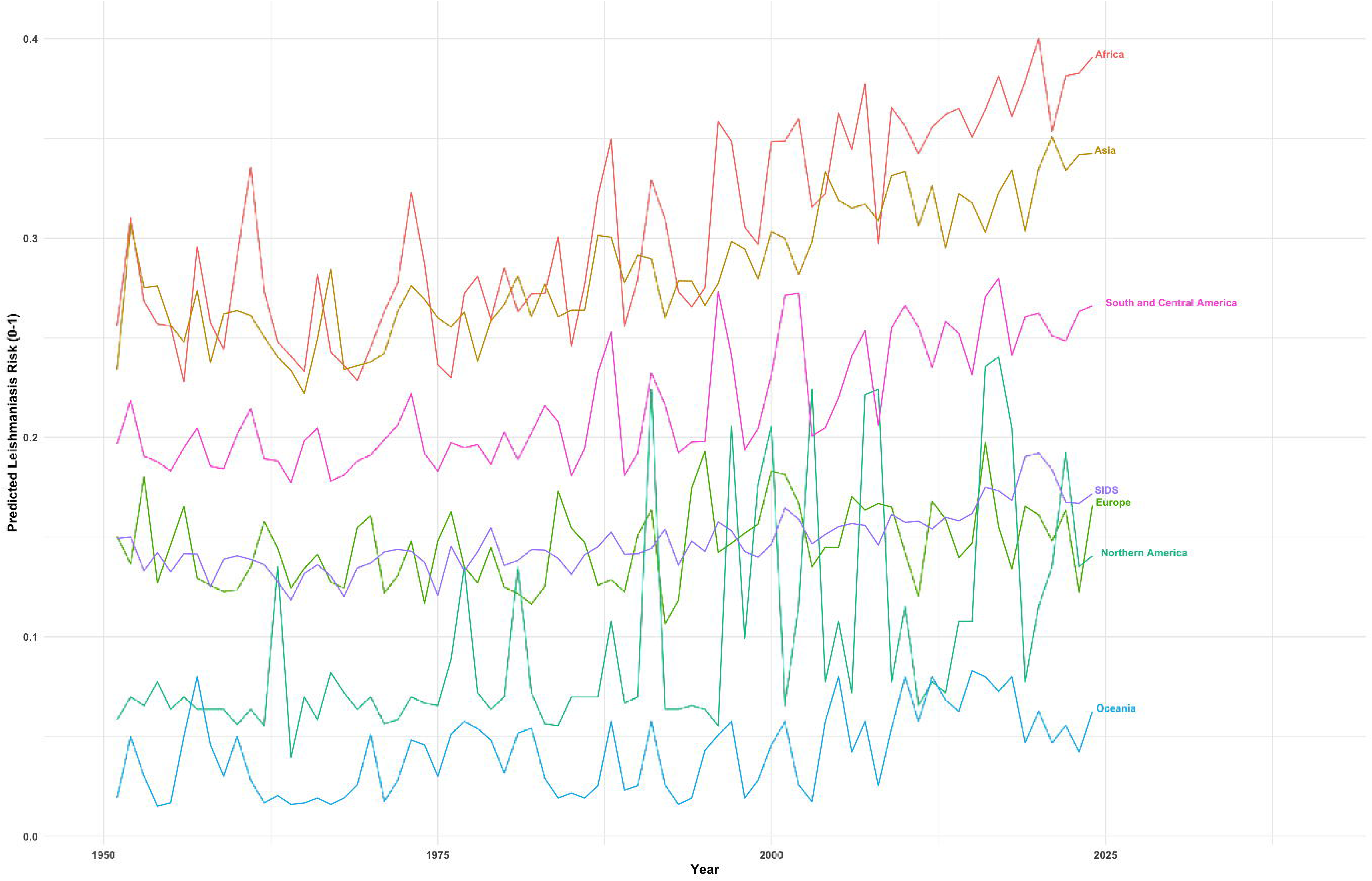
Time-series of predicted global annual leishmaniasis risk for 1951 to 2024 aggregated to the global regions (see **S3 Table** for region definitions) (**top panel**) and predicted European cutaneous (CL) and visceral (VL) leishmaniasis risk from 1970 to 2024 (**bottom panel**).

**Fig 5.**
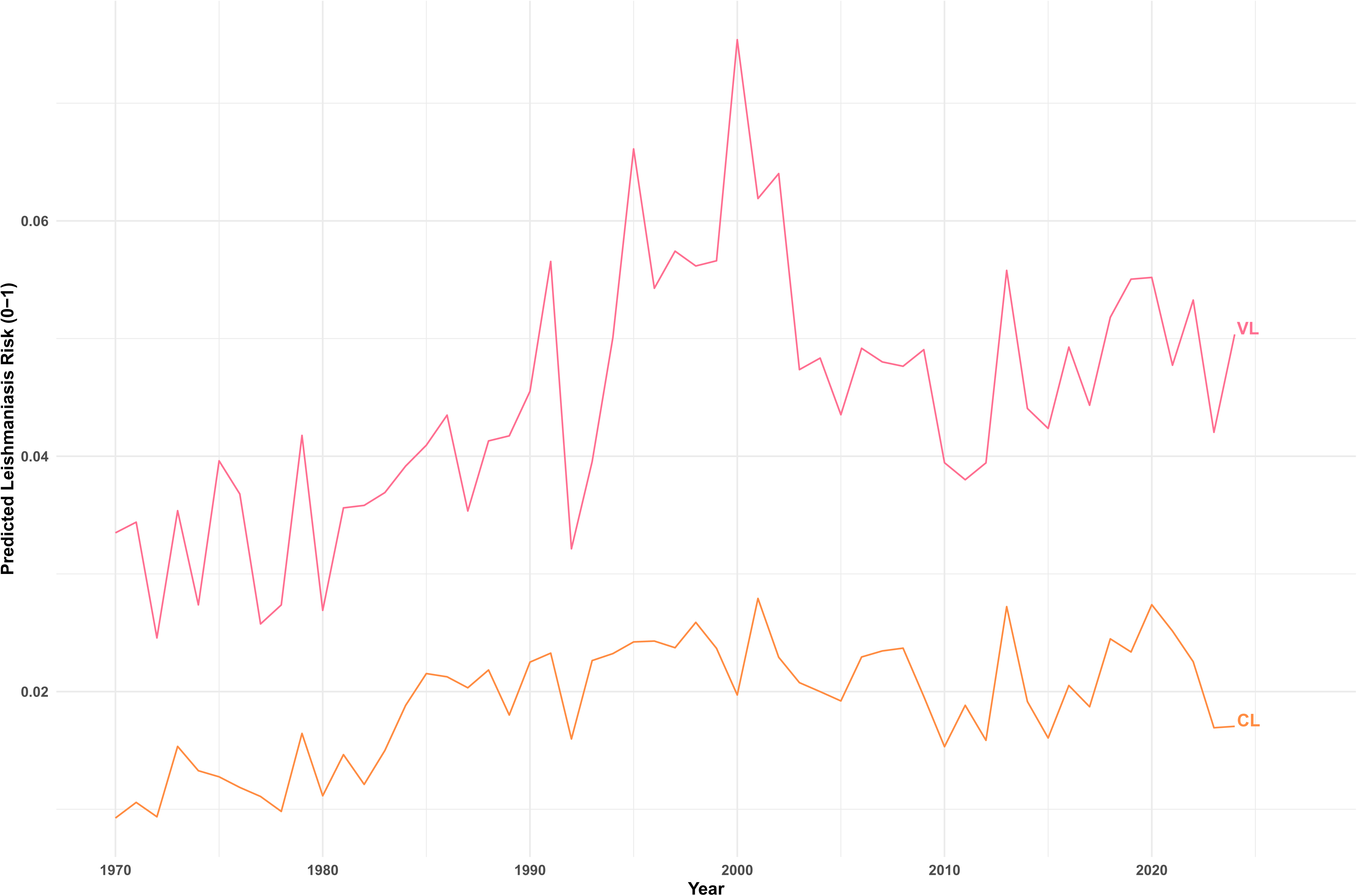
Change in predicted national leishmaniasis risk comparing the baseline and comparator periods in the global analysis (1951-1960 v 2014-2024) (**top panel**) and for cutaneous (CL) and visceral (VL) leishmaniasis in Europe by NUTS2 (1981-2010 v 2015-2024) (**bottom panel**).

In the Europe model predictions, despite only modest increases at the continental scale (1.28% for CL and 2.47% for VL for 1981-2010 v 2015-2024, harmonious with the global analysis) (**Fig 4**), leishmaniasis risk are highly spatially heterogeneous across Europe and appear strongly linked to inequity. When comparing risk from 1981-2010 and 2015-2024, the results suggest a northward expansion of VL, with an increase in VL risk of 29% in northern Europe, with some Nordic countries seeing significant increases (e.g., Finland (+41.7%) and Sweden (+58.2%). However, in the Nordic countries the absolute risk remains relatively low. Whereas, for CL, the risk appears to be increasing in southern and eastern Europe, finding a 24.6% increase in eastern Europe, and large predicted increases in risk in southern Spain, southern France, eastern Italy and southern regions of the Balkans and Greece, with some NUTS2 areas in these regions seeing a >100% increase in risk (**Fig 5**).

Across Europe, predicted sand fly risk has increased from 1981-2010 to 2015-2024 for *P. neglectus, P. ariasi* and *P. perniciosus* by 0.46, 7.49 and 4.12%, respectively, and decreased slightly for *P. perfiliewi* and *P. tobbi* by-1.08 and-0.41, respectively. These changes were highly spatially heterogeneous, with areas which were already endemic for sand flies having the smallest changes, and emerging settings, such as northern Europe, seeing large changes over the two time periods (see **Table 4** and sand fly endemic areas in **S2 Fig**).

**Table 4:**
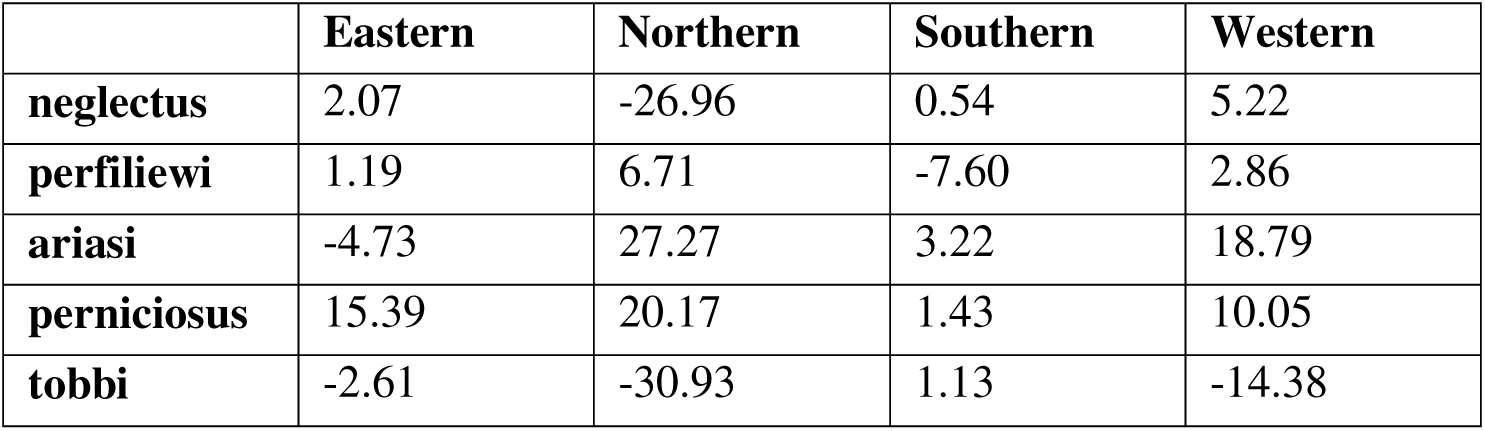
Change in predicted risk (%) for the five Phlebotomus sand flies known to transmit Leishmania in Europe by European region from 1981-2010 and 2015-2024 (see **S4 Table** for Europe region definitions)

|  | Eastern | Northern | Southern | Western |
| --- | --- | --- | --- | --- |
| **neglectus** | 2.07 | -26.96 | 0.54 | 5.22 |
| **perfliewi** | 1.19 | 6.71 | -7.60 | 2.86 |
| **ariasi** | -4.73 | 27.27 | 3.22 | 18.79 |
| **perniciosus** | 15.39 | 20.17 | 1.43 | 10.05 |
| **tobbi** | -2.61 | -30.93 | 1.13 | -14.38 |

## Discussion

The aim of the work presented here was to map the distribution of leishmaniasis based on the climatic suitability of the vector and socio-economic risk factors which lead to disease at both a global and regional (Europe) level. A machine learning approach was used, which can explore a large range of covariates and providing powerful predictions. Climatic indicators, land use cover, elevation and socio-economic indices were explored and trained on a range of models across several disease (CL, VL and leishmaniasis) and sand fly (5 Europe endemic species) binary outcomes. Land use features had the greatest SHAP values during covariate exploration, particularly land cover characterised as shrublands. Sandflies thrive in shady, humid and sheltered environments where they can rest during the day and maintain moisture, additionally, where they have readily available soil to lay eggs. Shrublands and forests are also a common habitat for several key hosts for sand flies including rodents and lagomorphs, providing ample sources for blood meals.^46,47^ Additional important environmental variables included mean temperature in the coldest month and humidity (in the warmest and driest quarters), potentially facilitating sand fly survival and extending activity seasons by reducing the number of months where minimum/maximum thresholds were met (e.g., during very hot and dry summers and/or during very cold winters).^48,49^

Socio-economic features proved to be powerful predictors at both the global and regional scale, supporting the evidence that leishmaniasis is a disease of inequity, even in more developed regions such as Europe. Poverty, poor access to healthcare, malnutrition and co-morbidities are global risk factors for leishmaniasis. These risk factors have been particularly pronounced during conflicts and in refugee camps in western Asia, namely Afghanistan and Syria,^50,51^ where ∼200,000 cases were reported in the early 2000s in Kabul alone, and ∼7,700 estimated cases per month in Syria and neighbouring Lebanon starting in around 2011. Risk factors of leishmaniases differ among European countries: VL is still primarily a paediatric disease in areas where poverty and malnutrition are more prevalent, particularly in eastern Europe. In western Europe, the majority of VL cases occur in immunosuppressed adults such as those with HIV comorbidity, those undergoing immunosuppressive therapies, and individuals with cancer.^19^ Inequalities in access to healthcare, social and economic burdens and stigma associated with leishmaniasis, whilst not as pronounced as in endemic low-income countries, are to be expected in the more marginalised and lower socioeconomic populations of Europe, including migrants and refugees.^52^ In future analysis it may be beneficial to explore a greater array of socio-economic covariates which may allow a greater understanding for why AROPE and HDI were powerful predictors of leishmaniasis here e.g., is this linked to income, healthcare, education etc.

Globally, leishmaniasis risk has increased by 17% since the 1990s, according to the model predictions here, which has also been found in other modelling studies, such as one global study which found that age-standardised prevalence has increased by ∼22.3%. However, Disability Adjusted Life Years for leishmaniasis globally has decreased according to the same study.^53^ Predicted risk for the most recent year of the analysis (2024) found Africa, Asia and South and Central America to have the highest values, particularly if Asia was further subset to the Eastern Mediterranean region. The results are harmonious with current known burden, with about 95% of CL cases occurring in the Americas, the Mediterranean basin and West and Central Asia and >90% of VL cases reported in Brazil, Ethiopia, India, Kenya, Somalia, South Sudan, and Sudan.^54^ These findings help support the model performance metrics, that these models are capable of accurately predicting leishmaniasis risk spatially. Africa was the region with the largest increase over the study period, largely due to high increases in risk in South Sudan, Guinea, Zambia and Namibia. These changes may be linked to changes in temperature, rainfall and humidity linked to climate change, along with land use changes including deforestation, irrigation, dam construction and urbanisation, which have all occurred in parallel to demographic pressures and poverty.^55,56^ However, there are severe data gaps in Africa, and therefore understanding the true burden continues to be a challange.^57^

For Europe, there was a slight projected increase in risk for leishmaniasis of 1.28% for CL and 2.47% for VL when comparing 1981-2010 and 2015-2024 at a continental scale. These results were harmonious with the global analysis, which also found modest increases across the continent. However, these results had high spatial heterogeneity especially at sub-national spatial resolutions. The leishmaniasis predictions largely match predicted changes to sand fly risk, which is reassuring that the model is performing correctly, as leishmaniasis should not have high predictions in areas with low sand fly predictions. The smallest changes in risk were found in areas already endemic for sand flies and the largest in non-endemic areas, suggesting that climatic suitability for sand flies is expanding across Europe, a finding also suggested in other studies.^43,58^ Additionally, the model predicted an expansion of warm-adapted species such as *P. ariasi, P. perniciosus* and *P. neglectus*, which favor higher temperature and extended warm seasons and a contraction of *P. perfiliewi* and *P. tobbi*, species which prefer cooler and more humid niches, finding that southern European summers are becoming too hot and dry.^59,60^ For leishmaniasis, the predictions suggest an expansion of endemic VL northward from the Mediterranean region, whereas, for emerging CL, the risk appears to be increasing in the more temperate areas of southern and eastern Europe, potentially exacerbated by their geographic proximity to endemic countries in western Asia and North Africa (although this is not accounted for in the model). This finding was supported by large increases in CL risk in southern Spain, southern France, eastern Italy and southern regions of the Balkans and Greece, known areas for refugee influxes due to conflicts and instability in western Asia and North Africa.^61,62^ These findings further highlight the need for refugees and asylum seekers to be adequately and efficiently processed and their health needs assessed, to ensure their health, reduce the impact of health systems, protect population health and the importation of emerging diseases, and to collate robust data to understand the scale of importations.

A major limitation of the approach used here is our inability to clearly distinguish between autochthonous and imported cases, which limits the interpretation of climatic suitability for disease transmission. In the case of an imported human or animal case, disease transmission might have occurred under different climatic conditions. Imported leishmaniasis cases are common due to movements associated with tourism, migration, and military operations.^63,64^ This includes imported infections in pets and the risk of working with and rescuing stray dogs which can establish transmission in non-endemic countries. Major obstacles to the control of leishmaniasis emergence in the EU and its neighbourhood countries include limitations associated with insufficient surveillance and control, lack of dedicated resources, and the perception that leishmaniases are regarded as a local, rather than a transnational problem.^65^ Furthermore, leishmaniasis is a neglected tropical disease for which relevant and standardised data at the global scale are scarce, resulting from differences in regional disease reporting and surveillance programmes. Spatial heterogeneity in reporting therefore resulted in likely spatial and temporal detection bias in our leishmaniasis dataset used here, both in training and testing. Human leishmaniasis is not a reportable disease in the European Union, and national notification is not compulsory in all endemic European countries.^65^ Even in those countries with compulsory notification systems, underreporting is common, as exemplified by the difference in the number of cases reported by each country to the WHO Global Health Observatory Data Repository,^1^ the number of hospital discharges published by the ECDC,^19,66^ and a recently published retrospective surveillance study in Europe.^64^

Leishmaniasis is a highly complex disease system, with multiple species of parasites, vectors and hosts that interact in space and time and with the environment to maintain a particular transmission cycle. The current version of the modelling approach does not consider species-specific information or long-term changes in land use. Long term land use datasets are unlikely to become available as they largely rely on satellite capabilities, which have greatly improved only over the most recent decades, however some datasets are available based on paleo-reconstructions and may be explored in later iterations. Furthermore, obtaining datasets on rodent and canine populations would be highly beneficial to improving the modelling approach used here, as these are likely key risk factors in maintaining leishmaniasis in the human transmission cycle. Despite these data limitations, I believe that the best available data were used for the analysis here, particularly given our ambitions to explore a global analysis.

This modelling approach and the results presented here provide a valuable evidence base to guide policymakers by highlighting regions with elevated probabilities of leishmaniasis occurrence. By integrating climatic, land use, and socio-economic covariates, the models underscore the multifactorial nature of disease risk and the importance of considering both environmental and societal determinants. They delineate areas at risk for the emergence and re-emergence of leishmaniasis that should be prioritised for integrated surveillance of vectors, reservoirs, and human cases. This is particularly relevant for regions bordering high-burden countries in Africa, where climatic suitability is expanding, and for Europe, where warm-adapted sand fly species are predicted to increase their range northward while others contract under hotter, drier conditions. In addition to vector surveillance, attention should be directed to infected pet dogs imported from endemic areas, which may act as reservoirs and seed local transmission cycles. Refugee and migrant populations also require special consideration, as they may both reflect and amplify socio-economic vulnerabilities linked to leishmaniasis. The findings reinforce the need for robust health monitoring in these groups and highlight the broader role of inequality in shaping disease outcomes, even within higher-income European settings. Addressing these data challenges outlined here will require stronger surveillance systems, harmonised data collection across countries, and interdisciplinary collaboration. Overall, leishmaniasis continues to be neglected despite its wide geographic footprint, severe health impacts, and capacity to expand under climate change. There is an urgent need for increased attention, sustained funding, and research investment at the global scale. Education and awareness campaigns targeted at both the public and health professionals are essential to reduce stigma, improve early diagnosis, and strengthen preparedness. Without such coordinated efforts, the ongoing expansion of sand flies’ climatic suitability and the persistence of socio-economic inequities will continue to place more people at risk of a disease that is preventable, yet potentially disfiguring and deadly.

## Data Availability

All data used in the analyses are open access and referenced and citated accordingly throughout the manuscript. Two open access GitHub repositories are available for all code used in the analyses, one specific to the European analysis (https://github.com/GinaCharnley/Leishmaniasis-Europe) and another for the global analysis (https://github.com/GinaCharnley/Leishmaniasis-Global).

## Acknowledgements

GECC acknowledges joint Centre funding The author the UK Medical Research Council and the Department for International Development [MR/R0156600/1]. The author thanks members of the *Lancet Countdown* consortium for general feedback on an earlier report that informed the broader research context, but not the analyses, interpretation, or manuscript preparation presented here.

## Author Contribution

GECC conceptualised the study, collected and harmonised the appropriate data, designed and ran the analysis, interpretated the results and finalised the manuscipt.

## Consent to Participate declaration

not applicable

## Ethics and Consent to Participate declarations

not applicable

## Supporting Information Captions

**S1 Fig. Spatial distribution of the leishmaniasis data used as the outcome variable for model fitting for the full dataset from 1970-2012**, with the **top panel** showing the global national distribution of both visceral and cutaneous leishmaniasis cases combined (grey = no data) and the **bottom panel** showing the points were cases were reported for either cutaneous leishmaniasis (left) and visceral leishmaniasis (right) in decadal time periods.

**S2 Fig. Spatial distribution of the sand fly presence** and absence/unavailable data for the main species known for transmitting *Leishmania infantum* in Europe according to ECDC’s VectorNet for 2024.

**S1 Table. Explanation of the nineteen annual WorldClim bioclimatic variables and the additional soil water and humidity variables used here**, derived from monthly Copernicus ERA5 data. A quarter is defined in three-month intervals, starting from January. Notation: *i* = month, *Tavg* = average temperature for month *i*, *PPT* = average precipitation for month, *VSW =* average volumetric soil water, *RH* = average relative humidity, *SD* = standard deviation.

**S3 Fig. Shapley Additive Explanations (SHAP) Importance values** for the global analysis in red and the European in blue against the seven potential sand fly (species names) and disease outcomes (vl = visceral leishmaniasis and cl = cutaneous leishmaniasis). Bioclimatic codes (BIOXX) are defined in **S1 Table**, hurs = relative humidity and sm = volumetric soil water.

**S4 Fig. Pearson correlation coefficient values** for the environmental features considered here in the best fit models for the fitting dataset for the global analysis in red and the European analysis in blue (disease outcomes in the top plot and fly outcomes in the bottom plot).

**S2 Table. The top 10 features and their clusters** according to a Pearson correlation coefficient threshold of 0.7 selected for formula combinations in each model for the global and European analysis.

**S3 Table. Definitions of the global regions**, following the Lancet region definitions.

**S4 Table. Definitions of the European regions**, following the United Nations region definitions.

---

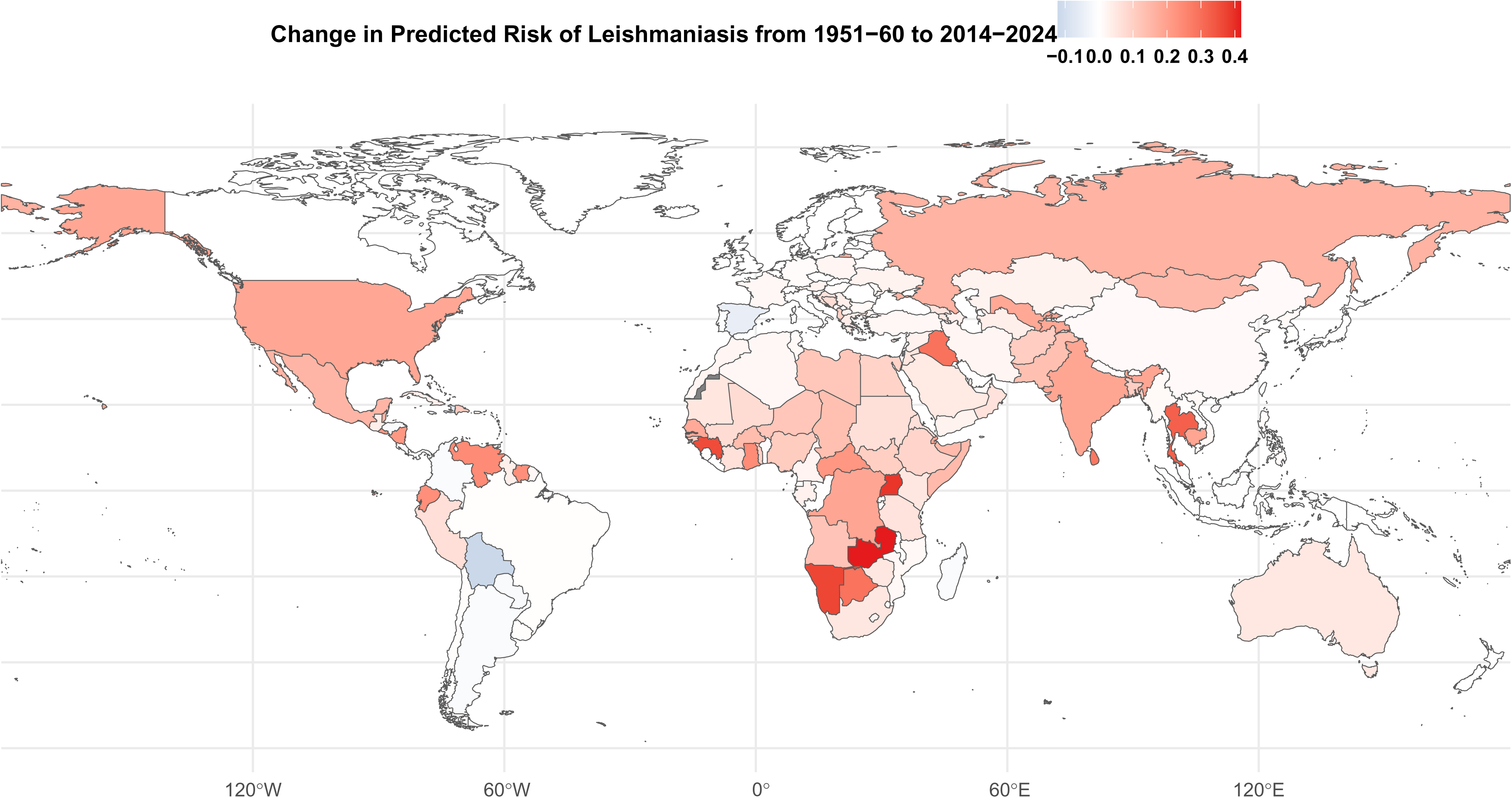

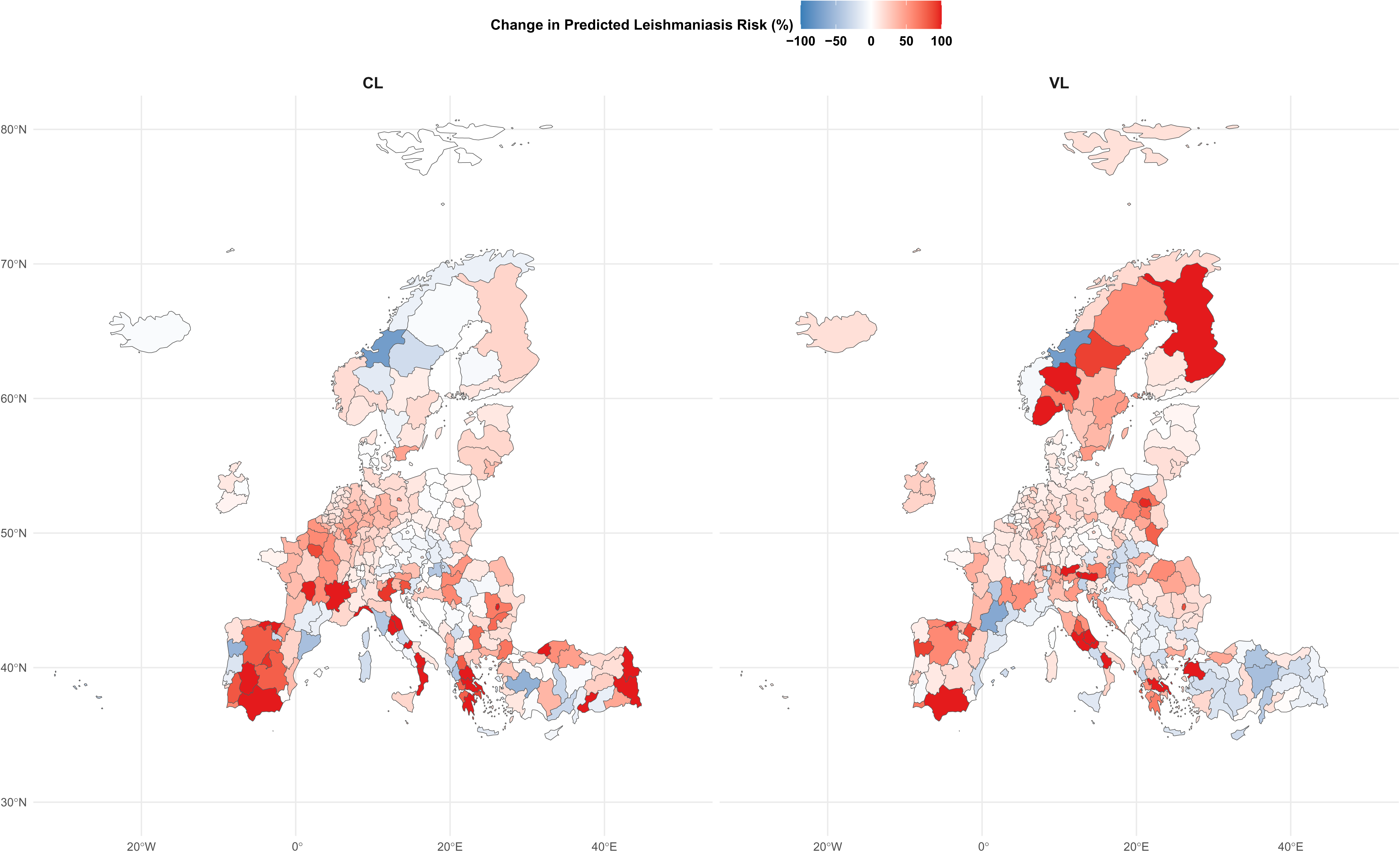

