## Supplementary material for "Climatic suitability for leishmaniasis at global and European scales": S1 Fig

**S1** **Fig**. **Spatial distribution of the leishmaniasis data used as the outcome variable for model fitting for the full dataset from 1970-2012**, with the **top panel** showing the global national distribution of both visceral and cutaneous leishmaniasis cases combined (grey = no data) and the **bottom panel** showing the points were cases were reported for either cutaneous leishmaniasis (left) and visceral leishmaniasis (right) in decadal time periods.
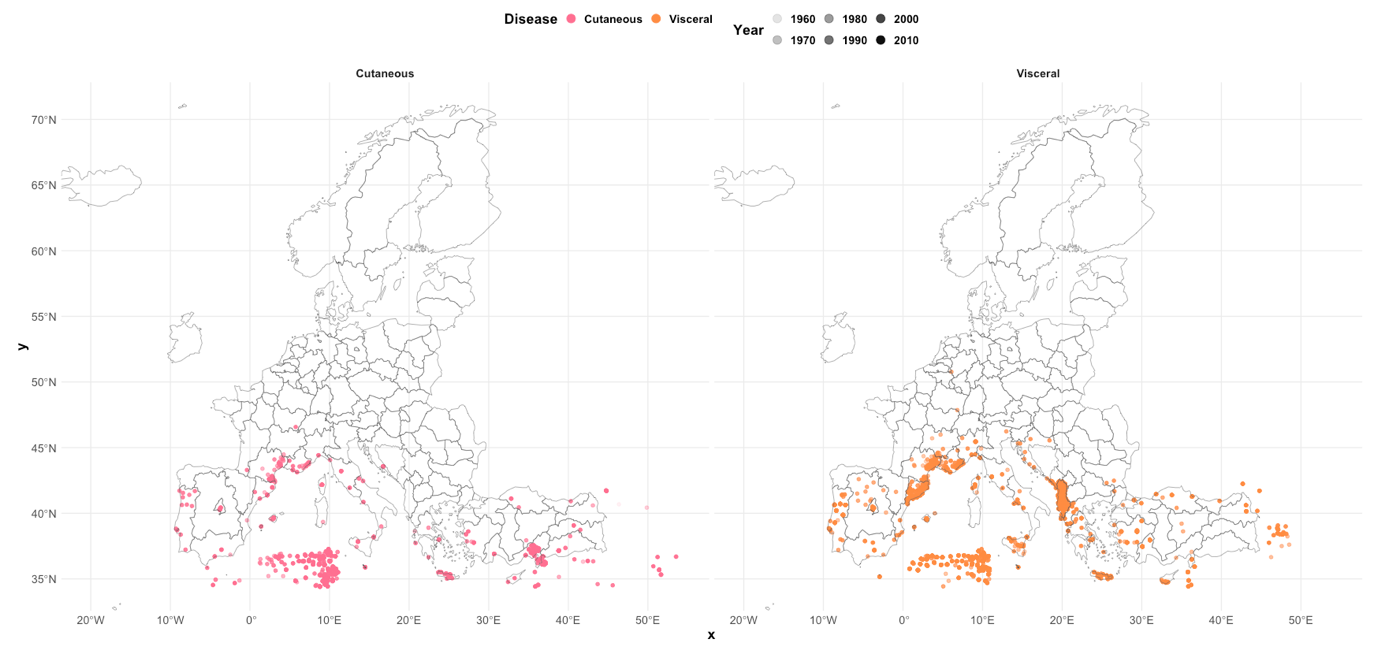

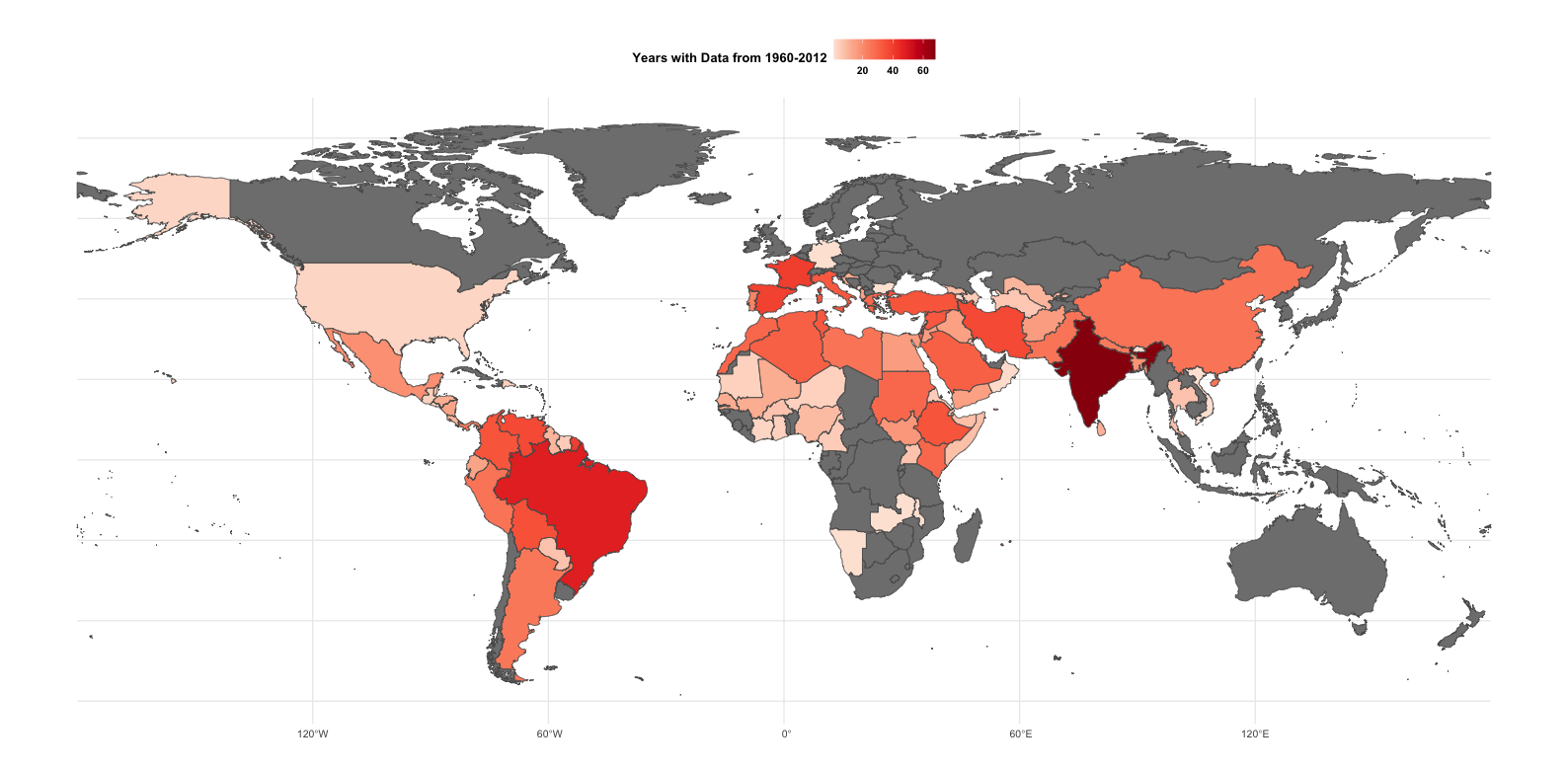

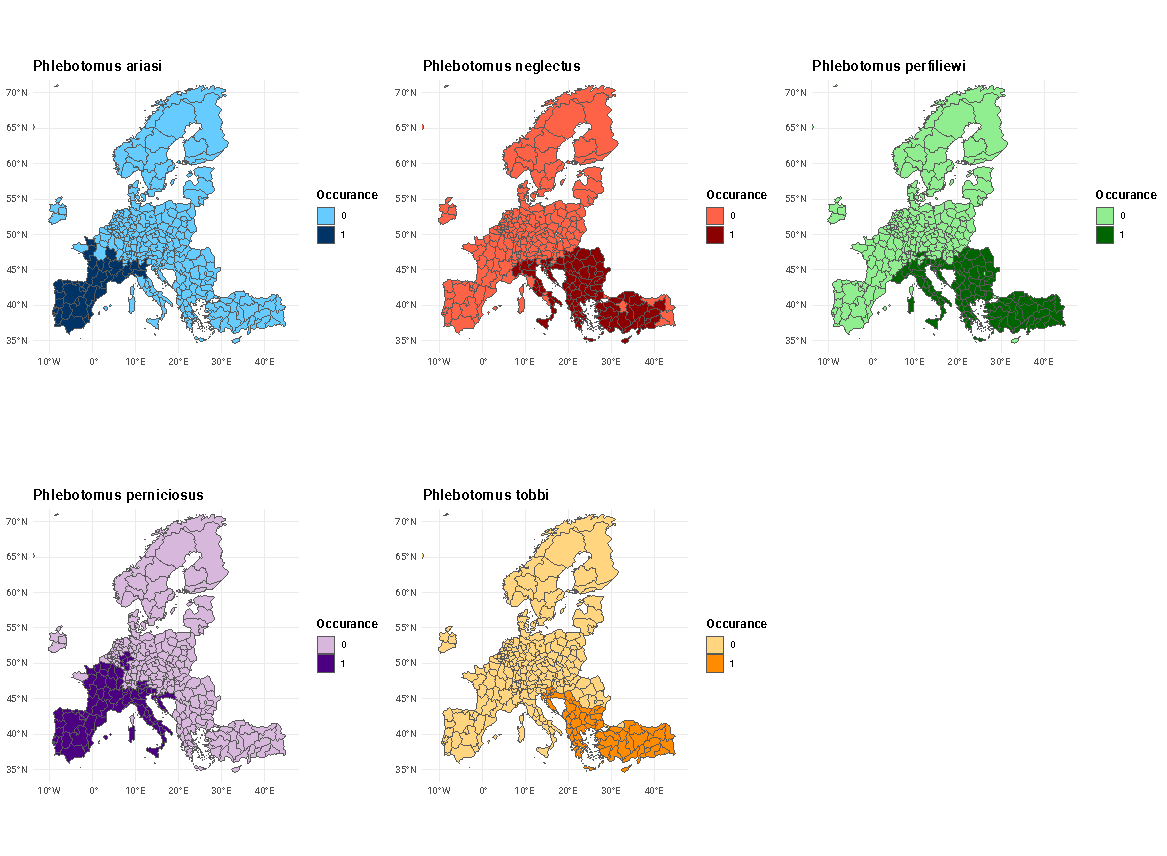

**S2 Fig. Spatial distribution of the sand fly presence** and absence/unavailable data for the main species known for transmitting *Leishmania infantum* in Europe according to ECDC's VectorNet for 2024.

**S1 Table. Explanation of the nineteen annual WorldClim bioclimatic variables and the additional soil water and humidity variables used here**, derived from monthly Copernicus ERA5 data. A quarter is defined in three-month intervals, starting from January. Notation: *i* = month, *Tavg* = average temperature for month *i*, *PPT* = average precipitation for month, *VSW* *=* average volumetric soil water, *RH* = average relative humidity, *SD* = standard deviation.

| **Indicator** | **Calculation** | **Unit** |
| --- | --- | --- |
| BIO1: Annual Mean Temperature | $BIO1= \frac{\Sigma_{i=1}^{i=12}{Tavg}_{i}}{12}$ | K to °C |
| BIO2: Mean Diurnal Range of Temperature | $BIO2= \frac{\Sigma_{i=1}^{i=12}({Tmax}_{i}-{Tmin}_{i})}{12}$ | K to °C |
| BIO3: Isothermality (Quantifies how large the day-to-night temperatures oscillate relative to the summer-to-winter oscillation) | $BIO3= \frac{BIO2}{BIO7}x100$ | % |
| BIO4: Temperature Seasonality  (The amount of temperature variation over a given year based on the standard deviation of monthly temperature averages) | $BIO4= SD\{{Tavg}_{1}\ldots\ldots,{Tavg}_{12}\}$ | K to °C (x100) |
| BIO5: Max Temperature of Warmest Month | $BIO5= max(\left\{ {Tavg}_{1}\ldots\ldots,{Tavg}_{12} \right\})$ | K to °C |
| BIO6: Min Temperature of Coldest Month | $BIO6= min(\left\{ {Tavg}_{1}\ldots\ldots,{Tavg}_{12} \right\})$ | K to °C |
| BIO7: Temperature Annual Range  (A measure of temperature variation over a year) | $BIO7= BIO5-BIO6$ | K to °C |
| BIO8: Mean Temperature of Wettest Quarter | $BIO8= \frac{\Sigma_{i=1}^{i=3}{Tavg}_{i}}{3}$  where *i* represents the quarter with the highest total precipitation | K to °C |
| BIO9: Mean Temperature of Driest Quarter | $BIO9= \frac{\Sigma_{i=1}^{i=3}{Tavg}_{i}}{3}$  where *i* represents the quarter with the lowest total precipitation | K to °C |
| BIO10: Mean Temperature of Warmest Quarter | $BIO10= \frac{\Sigma_{i=1}^{i=3}{Tavg}_{i}}{3}$  where *i* represents the quarter with the highest average temperature | K to °C |
| BIO11: Mean Temperature of Coldest Quarter | $BIO11= \frac{\Sigma_{i=1}^{i=3}{Tavg}_{i}}{3}$  where *i* represents the quarter with the lowest average temperature | K to °C |
| BIO12: Annual Precipitation  (Sum of all monthly precipitation values) | $BIO12= \sum_{i=1}^{i=12} {PPT}_{i}$ | mm |
| BIO13: Precipitation of Wettest Month | $BIO13= max(\left\{ {PPT}_{1}\ldots\ldots,{PPT}_{12} \right\})$ | mm |
| BIO14: Precipitation of Driest Month | $BIO14= max(\left\{ {PPT}_{1}\ldots\ldots,{PPT}_{12} \right\})$ | mm |
| BIO15: Precipitation Seasonality  (A measure of the variation in monthly precipitation totals over the course of the year: coefficient of variation) | $BIO15= \frac{SD\{{PPT}_{1}\ldots\ldots,{PPT}_{12}\}}{1+(\frac{BIO12}{12})}x100$ | Dimensionless |
| BIO16: Precipitation of Wettest Quarter | $BIO16= \frac{\Sigma_{i=1}^{i=3}{PPT}_{i}}{3}$  where *i* represents the quarter with the highest total precipitation | mm |
| BIO17: Precipitation of Driest Quarter | $BIO17= \frac{\Sigma_{i=1}^{i=3}{PPT}_{i}}{3}$  where *i* represents the quarter with the lowest total precipitation | mm |
| BIO18: Precipitation of Warmest Quarter | $BIO18= \frac{\Sigma_{i=1}^{i=3}{PPT}_{i}}{3}$  where *i* represents the quarter with the highest average temperature | mm |
| BIO19: Precipitation of Coldest Quarter | $BIO19= \frac{\Sigma_{i=1}^{i=3}{PPT}_{i}}{3}$  where *i* represents the quarter with the lowest average temperature | mm |
| Mean Volumetric Soil Water of Wettest Quarter | ${VSW}_{wet}= \frac{\Sigma_{i=1}^{i=3}{PPT}_{i}}{3}$  where *i* represents the quarter with the highest total precipitation | m^3^ m^-3^ |
| Mean Volumetric Soil Water of Driest Quarter | ${VSW}_{dry}= \frac{\Sigma_{i=1}^{i=3}{PPT}_{i}}{3}$  where *i* represents the quarter with the lowest total precipitation | m^3^ m^-3^ |
| Mean Volumetric Soil Water of Warmest Quarter | ${VSW}_{warm}= \frac{\Sigma_{i=1}^{i=3}{PPT}_{i}}{3}$  where *i* represents the quarter with the highest average temperature | m^3^ m^-3^ |
| Mean Volumetric Soil Water of Coldest Quarter | ${VSW}_{cold}= \frac{\Sigma_{i=1}^{i=3}{PPT}_{i}}{3}$  where *i* represents the quarter with the lowest average temperature | m^3^ m^-3^ |
| Mean Relative Humidity of Wettest Quarter | ${RH}_{wet}= \frac{\Sigma_{i=1}^{i=3}{PPT}_{i}}{3}$  where *i* represents the quarter with the highest total precipitation | % |
| Mean Relative Humidity of Driest Quarter | ${RH}_{dry}= \frac{\Sigma_{i=1}^{i=3}{PPT}_{i}}{3}$  where *i* represents the quarter with the lowest total precipitation | % |
| Mean Relative Humidity of Warmest Quarter | ${RH}_{warm}= \frac{\Sigma_{i=1}^{i=3}{PPT}_{i}}{3}$  where *i* represents the quarter with the highest average temperature | % |
| Mean Relative Humidity of Coldest Quarter | ${RH}_{cold}= \frac{\Sigma_{i=1}^{i=3}{PPT}_{i}}{3}$  where *i* represents the quarter with the lowest average temperature | % |

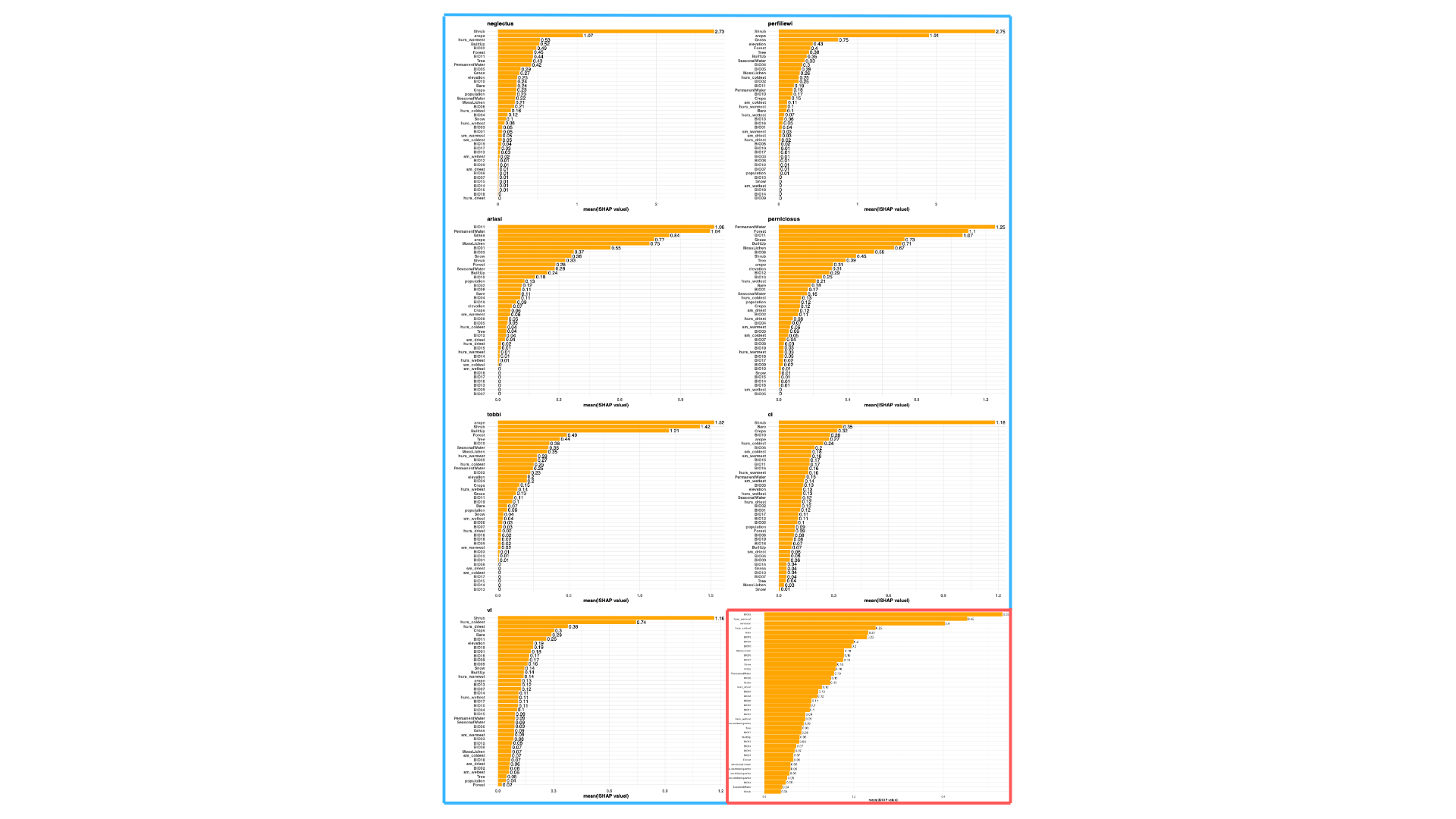
**S3 Fig. Shapley Additive Explanations (SHAP) Importance values** for the global analysis in red and the European in blue against the seven potential sand fly (species names) and disease outcomes (vl = visceral leishmaniasis and cl = cutaneous leishmaniasis). Bioclimatic codes (BIOXX) are defined in **S1 Table**, hurs = relative humidity and sm = volumetric soil water.

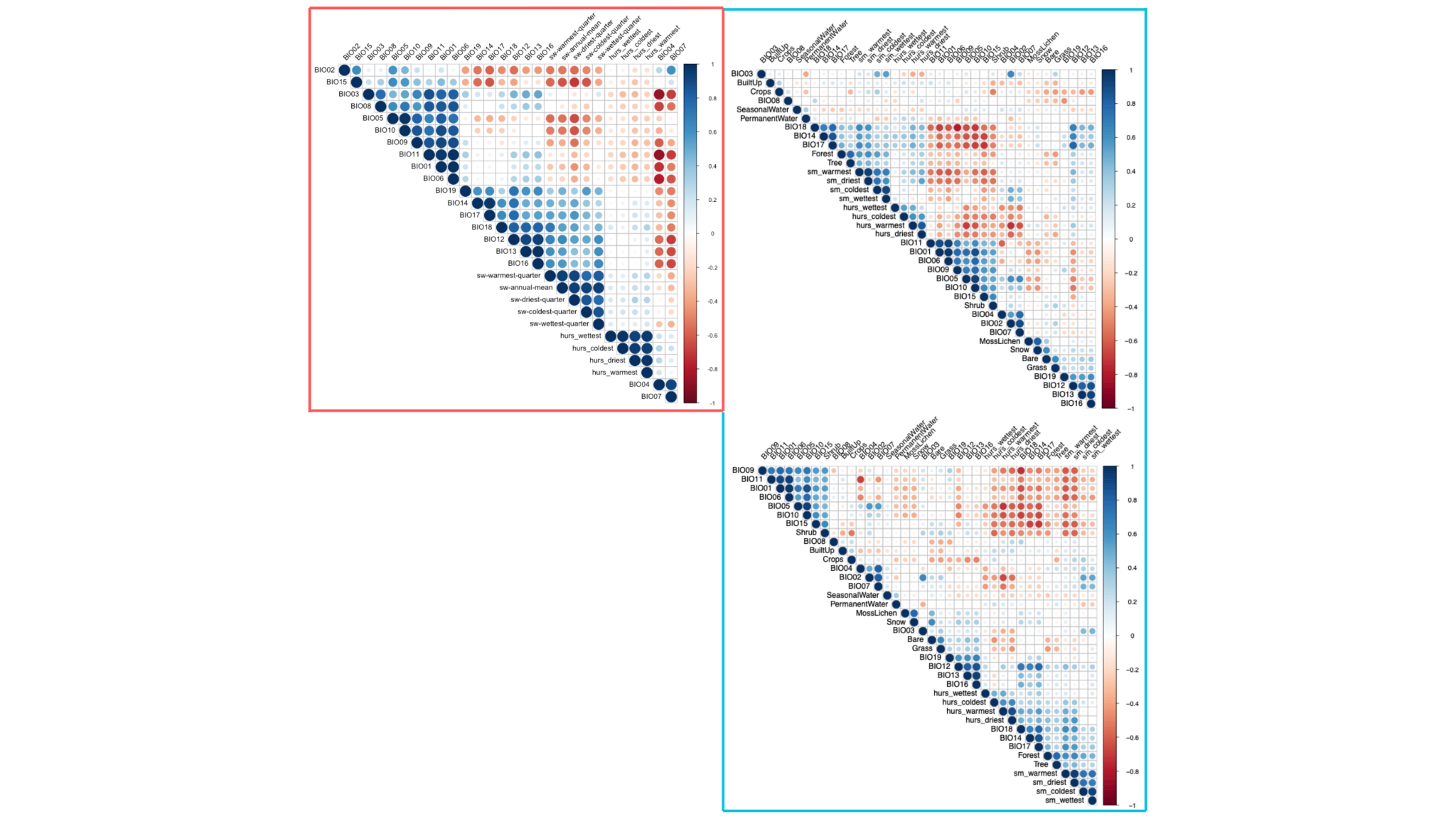

**S4 Fig. Pearson correlation coefficient values** for the environmental features considered here in the best fit models for the fitting dataset for the global analysis in red and the European analysis in blue (disease outcomes in the top plot and fly outcomes in the bottom plot).

**S2 Table. The top 10 features and their clusters** according to a Pearson correlation coefficient threshold of 0.7 selected for formula combinations in each model for the global and European analysis.

| **Feature** | **Region** | **Outcome** | **Cluster** |
| --- | --- | --- | --- |
| **BIO11** | European | ariasi | 1 |
| **PermanentWater** | European | ariasi | 2 |
| **Grass** | European | ariasi | 3 |
| **arope** | European | ariasi | 4 |
| **MossLichen** | European | ariasi | 5 |
| **BIO01** | European | ariasi | 1 |
| **BIO03** | European | ariasi | 6 |
| **Snow** | European | ariasi | 5 |
| **BIO10** | European | ariasi | 1 |
| **BIO02** | European | ariasi | 7 |
| **Shrub** | European | cl | 1 |
| **Bare** | European | cl | 2 |
| **Crops** | European | cl | 3 |
| **BIO10** | European | cl | 4 |
| **arope** | European | cl | 5 |
| **hurs_coldest** | European | cl | 6 |
| **BIO06** | European | cl | 4 |
| **sm_coldest** | European | cl | 7 |
| **sm_warmest** | European | cl | 7 |
| **PermanentWater** | European | cl | 8 |
| **Shrub** | European | neglectus | 1 |
| **arope** | European | neglectus | 2 |
| **hurs_warmest** | European | neglectus | 3 |
| **BuiltUp** | European | neglectus | 4 |
| **BIO02** | European | neglectus | 3 |
| **Forest** | European | neglectus | 5 |
| **BIO11** | European | neglectus | 6 |
| **Tree** | European | neglectus | 5 |
| **BIO05** | European | neglectus | 7 |
| **BIO10** | European | neglectus | 7 |
| **Shrub** | European | perfiliewi | 1 |
| **arope** | European | perfiliewi | 2 |
| **Grass** | European | perfiliewi | 3 |
| **elevation** | European | perfiliewi | 4 |
| **Forest** | European | perfiliewi | 5 |
| **BIO04** | European | perfiliewi | 6 |
| **BIO05** | European | perfiliewi | 7 |
| **hurs_coldest** | European | perfiliewi | 8 |
| **BIO02** | European | perfiliewi | 9 |
| **BIO11** | European | perfiliewi | 10 |
| **PermanentWater** | European | perniciosus | 1 |
| **Forest** | European | perniciosus | 2 |
| **BIO11** | European | perniciosus | 3 |
| **Grass** | European | perniciosus | 4 |
| **BuiltUp** | European | perniciosus | 5 |
| **MossLichen** | European | perniciosus | 6 |
| **BIO06** | European | perniciosus | 7 |
| **BIO12** | European | perniciosus | 8 |
| **BIO13** | European | perniciosus | 8 |
| **hurs_wettest** | European | perniciosus | 9 |
| **arope** | European | tobbi | 1 |
| **Shrub** | European | tobbi | 2 |
| **BuiltUp** | European | tobbi | 3 |
| **Forest** | European | tobbi | 4 |
| **Tree** | European | tobbi | 4 |
| **BIO10** | European | tobbi | 5 |
| **hurs_warmest** | European | tobbi | 6 |
| **BIO05** | European | tobbi | 5 |
| **hurs_coldest** | European | tobbi | 7 |
| **BIO02** | European | tobbi | 8 |
| **Shrub** | European | vl | 1 |
| **hurs_coldest** | European | vl | 2 |
| **hurs_driest** | European | vl | 3 |
| **Crops** | European | vl | 4 |
| **Bare** | European | vl | 5 |
| **BIO11** | European | vl | 6 |
| **elevation** | European | vl | 7 |
| **BIO19** | European | vl | 8 |
| **BIO01** | European | vl | 6 |
| **Snow** | European | vl | 9 |
| **BIO02** | Global | leishmaniasis | 1 |
| **hurs_warmest** | Global | leishmaniasis | 2 |
| **elevation** | Global | leishmaniasis | 3 |
| **hurs_coldest** | Global | leishmaniasis | 2 |
| **Bare** | Global | leishmaniasis | 4 |
| **BIO19** | Global | leishmaniasis | 5 |
| **BIO09** | Global | leishmaniasis | 6 |
| **BIO10** | Global | leishmaniasis | 6 |
| **MossLichen** | Global | leishmaniasis | 7 |
| **BIO05** | Global | leishmaniasis | 6 |

**S3 Table. Definitions of the global regions**, following the Lancet region definitions.

| **Region** | **Country** |
| --- | --- |
| Europe | Switzerland, Norway, Iceland, Denmark, Sweden, Germany, Ireland, Netherlands, Belgium, Finland, Liechtenstein, United Kingdom, Luxembourg, Austria, Slovenia, Malta, Spain, France, Cyprus, Italy, Estonia, Czechia, Greece, Andorra, Poland, Latvia, Lithuania, Croatia, Portugal, San Marino, Slovakia, Hungary, Montenegro, Romania, Russian Federation, Serbia, Belarus, Bulgaria, Albania, Bosnia and Herzegovina, North Macedonia, Moldova (Republic of), Ukraine |
| Asia | Hong Kong, China (SAR), Singapore, Korea (Republic of), Japan, Israel, Türkiye, United Arab Emirates, Bahrain, Qatar, Saudi Arabia, Kuwait, Brunei Darussalam, Oman, Georgia, Afghanistan, Yemen, Malaysia, Thailand, Kazakhstan, China, Armenia, Iran (Islamic Republic of), Sri Lanka, Azerbaijan, Turkmenistan, Mongolia, Jordan, Uzbekistan, Viet Nam, Lebanon, Palestine, State of, Indonesia, Philippines, Kyrgyzstan, Bhutan, Tajikistan, Iraq, Bangladesh, India, Lao People's Democratic Republic, Myanmar, Nepal, Cambodia, Syrian Arab Republic, Pakistan |
| Oceania | Australia, New Zealand, Tonga |
| Northern America | Canada, United States |
| South and Central America | Chile, Argentina, Uruguay, Panama, Costa Rica, Mexico, Ecuador, Peru, Brazil, Columbia, Paraguay, Venezuela (Bolivarian Republic of), Bolivia (Plurinational State of), El Salvador, Nicaragua, Guatemala |
| SIDS | Saint Kitts and Nevis, Antigua and Barbuda, Bahamas, Trinidad and Tobago, Barbados, Seychelles, Palau, Mauritius, Grenada, Saint Vincent and the Grenadines, Dominican Republic, Cuba, Maldives, Guyana, Dominica, Marshall Islands, Fiji, Saint Lucia, Jamaica, Somoa, Belize, Nauru, Suriname, Cabo Verde, Tuvalu, Micronesia (Federated States of), Kiribati, Honduras, Vanuatu, Sao Tome and Principe, Comoros, Papua New Guinea, Timor-Leste, Solomon Islands, Haiti |
| Africa | Côte d'Ivoire, Tanzania (United Republic of), Lesotho, Senegal, Sudan, Djibouti, Malawi, Benin, Gambia, Eritrea, Ethiopia, Liberia, Madagascar, Guinea-Bissau, Congo (Democratic Republic of the), Guinea, Mozambique, Sierra Leone, Burkina Faso, Burundi, Chad, Mali, Niger, Central African Republic, South Sudan, Somalia, Libya, Algeria, Togo, Egypt, Morocco, South Africa, Botswana, Gabon, Equatorial Guinea, Eswatini (Kingdom of), Namibia, Ghana, Kenya, Congo, Angola, Cameroon, Zambia, Uganda, Zimbabwe, Nigeria, Rwanda, Mauritania |

**S4 Table.** **Definitions of the European regions**, following the United Nations region definitions.

| **Region** | **Country** |
| --- | --- |
| Western | Switzerland, Germany, Netherlands, Belgium, Liechtenstein, Luxembourg, Austria, France, Monaco |
| Northern | Norway, Iceland, Denmark, Sweden, Ireland, Finland, United Kingdom, Estonia, Latvia, Lithuania |
| Southern | Slovenia, Slovenia, Malta, Spain, Cyprus, Italy, Greece, Andorra, Croatia, Portugal, San Marino, Türkiye, Montenegro, Serbia, Albania, Bosnia and Herzegovina, North Macedonia |
| Eastern | Czechia, Poland, Slovakia, Hungary, Romania, Russian Federation, Belarus, Bulgaria, Moldova (Republic of), Ukraine |
